# Loss of *GFAP* causes optico-retinal dysplasia and vision impairment

**DOI:** 10.1101/2022.11.09.22282105

**Authors:** Menachem Viktor Khamo Sarusie, Cecilia Rönnbäck, Cathrine Jespersgaard, Sif Baungaard, Yeasmeen Ali, Line Kessel, Søren Tvorup Christensen, Karen Brøndum- Nielsen, Kjeld Møllgård, Thomas Rosenberg, Lars Allan Larsen, Karen Grønskov

## Abstract

Diagnosis of genetic diseases has taken tremendous steps forward since the human genome project and technical advancements such as next generation sequencing. However, in the past years it has become evident that the classical “one gene – one phenotype” model is insufficient to encompass the intricacies of human genetics. Examples are emerging that variants in a gene can cause quite diverging phenotypes depending on the specific location in the gene or on the specific type of variant. In the era of precision medicine this is important knowledge, both when interpreting genomic data, but also when designing treatment strategies.

Gain-of-function variants in *GFAP* leads to protein aggregation and is the cause of the severe neurodegenerative disorder Alexander Disease (AxD), while loss of GFAP function has been considered benign. Here, we report a loss-of-function variant in *GFAP* as the cause of optico-retinal dysplasia and vision impairment in a six-generation family.

Whole genome sequencing analysis of family members with gliosis of the optic nerve head and visual impairment revealed a frameshift variant in *GFAP* (c.928dup, p.(Met310Asnfs*113)) segregating with disease. Analysis of human embryonic tissues revealed strong expression of GFAP in retinal neural progenitors. A zebrafish model verified that c.928dup does not result in extensive GFAP protein aggregation and zebrafish *gfap* loss-of-function mutants showed vision impairment and retinal dysplasia, characterized by a significant loss of Müller glia cells and photoreceptor cells.

Our findings show how different mutational mechanisms can cause diverging phenotypes and reveal a novel function of GFAP in human eye development.

## INTRODUCTION

Glial Fibrillary Acidic Protein (GFAP) is an intermediate filament protein, expressed in astrocytes of the adult central nervous system (CNS) and retina of mammals^1,2^. In the developing human brain, GFAP is expressed in radial glial cells (RGCs) from 6 weeks post conception (wpc) and in astrocytes from 14 wpc^3^. The expression of GFAP in the developing retina is less well studied. Early immunohistochemical analyses did not reveal GFAP expression in retinal tissue of mouse embryos (E12-E20) ^4^, while a more recent study, based on embryos obtained from *S. canicula* (small-spotted catshark), identified GFAP expression in RNPs at early developmental stages and in Müller glia cells (MGCs) at later stages^5^. However, upregulation of GFAP expression in MGCs occurs in all vertebrate species in response to eye injury and disease. In fish and chicken, MGCs serve as a source for neural progenitors including photoreceptors and a scaffold for the progenitors to migrate upon to regenerating retina layers ^6–10^. Despite expression of GFAP in RNPs and MGCs in some species, the role of GFAP in retina development remains poorly understood.

Rare missense variants within *GFAP* cause Alexander disease (AxD, OMIM #203450) - an autosomal dominant inherited severe leukodystrophy with variable onset ranging from infant to adult life^11^. AxD is characterized by Rosenthal fibers (RFs), which are dense inclusions within astrocytes of the forebrain and/or hindbrain, caused by aggregation and accumulation of GFAP^12,13^. The majority of *GFAP* variants identified in AxD patients are *de novo* heterozygous missense variants, while a few cases with variants leading to in-frame protein alterations have been reported^13^. A single patient, carrying a *GFAP* nonsense variant (p.E312*), with a mild form of AxD has been reported^14^. The apparent absence of loss-of-function *GFAP* variants in patients, combined with supporting evidence from mouse models, suggest that AxD is caused by GFAP toxicity through a gain-of-function mechanism^12,15^. Recent research in rodent models of AxD proposes that suppression of GFAP expression, through *Gfap*-targeted antisense oligonucleotides, is a promising therapeutic approach to reverse the disease progression in AxD^16,17^. With these exciting and encouraging results in mind, it is important to delineate the normal function of GFAP and any possible side effects from suppression of GFAP.

Here, we report the discovery of a rare form of autosomal dominant optico-retinal disorder segregating in a six-generation Danish family. Genetic analysis led to identification of a heterozygous frameshift variant of *GFAP* segregating with the disease in the family.

Analysis of human embryonic tissues and zebrafish *gfap* mutants confirmed that GFAP plays an important role in retinal development and that loss of GFAP can cause visual impairment.

## RESULTS

### Identification of a family with inherited dysplasia of the optic nerve head with retinal involvement

A six-generation Danish family with low vision was discovered and followed during a period of 35 years (Fig. 1a). Clinical examination was performed in seven family members (IV-1, IV-6, V-1, V-3, V-7, VI-2, and VI-4) with best corrected visual acuity (BCVA) from normal to low vision. Table S1 summarizes clinical and genetic data for four individuals (IV-6 (non-penetrant), V-1, V-3, and V-7) and figure 1b shows OCT of three individuals (V-1, V-3, and V-7) and optic nerve head photographs of all seven individuals.

**Figure 1.**
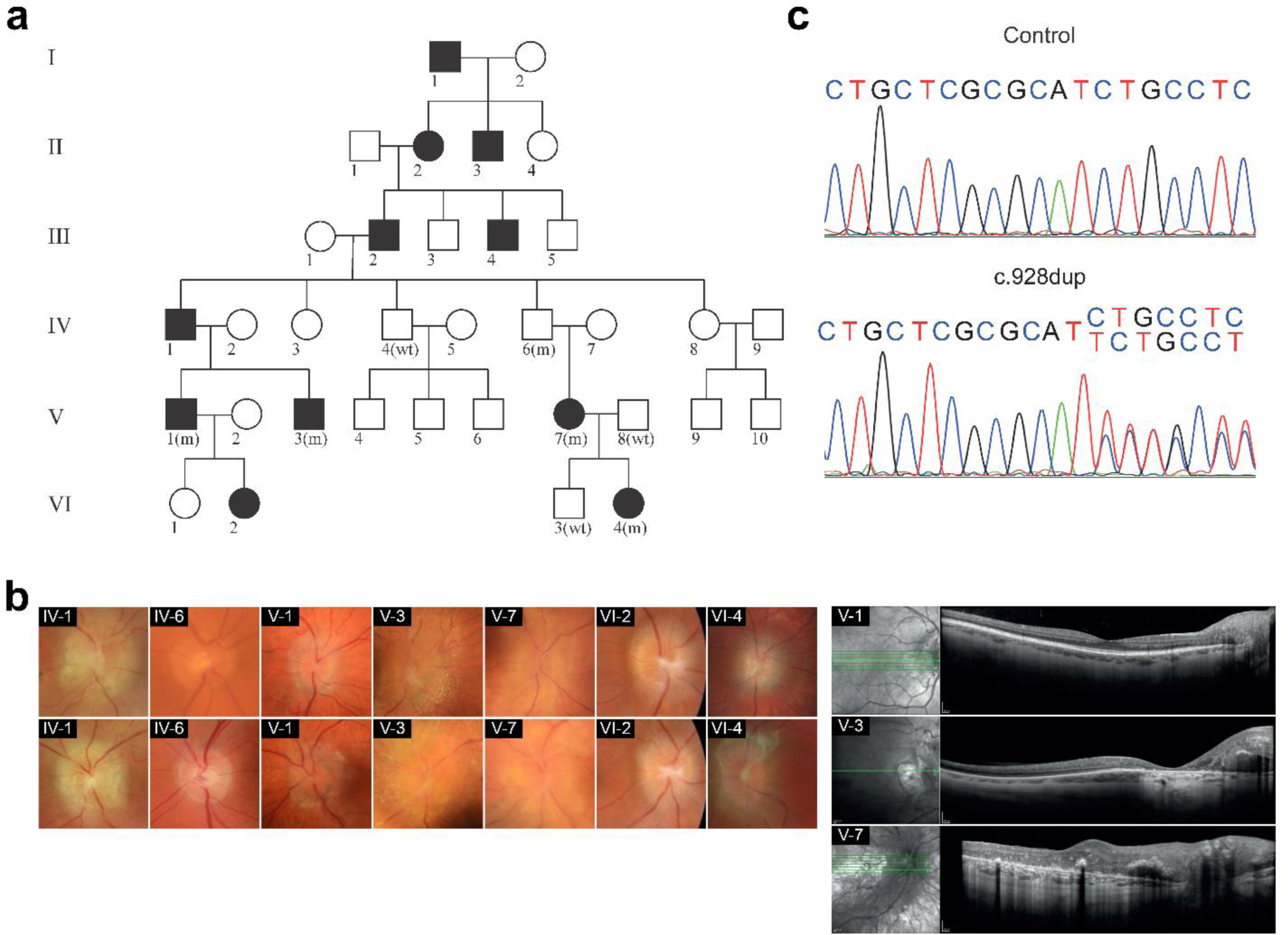
Mutation of GFAP is associated with retinal dysplasia and vision loss. **a.** Pedigree of a family with an autosomal dominant optical dysplasia with retinal involvement and vision loss. Affected individuals are shown with black fill symbols and unaffected with open symbols. The affected individuals V-1, V-3, V-7, VI-4 and an obligate carrier IV-6 were heterozygous for a c.928dup variant (m) in *GFAP*, while three unaffected individuals (IV-4, V-8 and VI-3) were found not to be carriers (wt). **b.** Clinical data. The fundoscopic phenotype showed varied degrees of gliosis of the optic nerve head resulting in a dome-shaped pellucid epiretinal veil and blurring of the disc margins (left panel, showing right and left eye). Subretinal fluid was observed. Three of the affected individuals (IV-1, V-3, and V-7) presented a massive glia-like mass covering the optic disc resembling pseudopapilledema as shown by Macular- and optic nerve OCT (Right panel). **c.** DNA sequence of *GFAP* nucleotide c.916-934 from a healthy (control) and an affected (c.928dup) individual.

Three family members with macular involvement (IV-1, V-3 and V-7) were visually impaired (20/60 and 20/120 at latest follow-up) whereas vision was normal or slightly subnormal in family members without macular involvement. The earliest examinations of family members were carried out at age six and seven years. The condition was largely stationary in individuals followed through decades.

Noticeable clinical heterogeneity was present. The fundoscopic phenotype showed varied degrees of gliosis of the optic nerve head resulting in a dome-shaped pellucid epiretinal veil and blurring of the disc margins. Three of the affected individuals (IV-1, V-3, and V-7) presented a massive glia-like mass covering the optic disc resembling pseudopapilledema. The mass extended along both arteries and veins, which demonstrated sheating/gliosis, a cork-shrew tortuous appearance, and punctuate and splinter hemorrhages. Subretinal fluid was observed. The papillo-macular area including the fovea showed atrophy of the pigment epithelium (Fig. 1b, Table S1). No retinal dragging was observed in any of the affected individuals. Individuals V-3 and V-7 with macular atrophy had central scotomas as measured with Goldmann perimetry; color vision and dark- and light-adapted full-field ERG, however, were normal. OCT confirmed the abnormal morphology of the optic disc in individuals V-3 and V-7 with the epiretinal glial-like mass extended temporally to the macular region, which showed disorganized inner retinal layering (Fig. 1b). The tissue complex measured as much as 1.3 mm in height in the subject with the most excessive glial growth. In the fovea of individuals V-3 and V-7 the retina was disorganized with clumped hyperpigmentations and atrophic retinal pigment epithelium (Fig. 1b), which probably explains the poor vision. Fluorescein angiography of individual V-7 at the age of 18 showed that the epi-papillary glial mass was richly vascularized, which continued into the papillo-macular area following the nerve fiber pattern (not shown). Early and late profuse capillary leakage was present at the posterior pole and in the foveal area. All the examined individuals were in good health without signs of any systemic disease.

### Identification of a rare variant in the *GFAP* gene

Whole genome sequencing (WGS) with subsequent filtering of variants showed that three affected individuals (V-1, V-3 and V-7) were all heterozygous for five rare coding variants, which were not present in an unaffected individual (IV-4) (Fig. 1a, Table S2). Two of the five variants were synonymous (PEMT p.Pro19= and OR10H2 p.Thr116=), not predicted to affect splicing using the spice predictors in Alamut Visual, and were not further analysed. Neither *PEMT* nor *OR10H2* are associated with a human phenotype. Two variants were missense variants (GAS2L2 p.Pro288Leu and SOST p.Ala28Val). Biallelic variants in *GAS2L2* are associated with primary ciliary dyskinesia (OMIM 618449)^18^; GAS2L2 p.Pro288Leu was classified as VUS (variant of unknown significance) following ACMG guidelines^19^ and ClinGenome webpage^20^. Monoallelic variants in *SOST* are associated with Craniodiaphyseal dysplasia (OMIM 122860)^21^ and biallelic variants are associated with Sclerosteosis 1 (OMIM 269500)^22^. According to ACMG guidelines, the variant is classified as VUS, but the presence of 19 alleles out of 251433 in gnomAD v2.1.1 database, would be considered too high for a severe dominant disease with onset in childhood. We therefore considered it unlikely that these four variants were the cause of retinal dysplasia in the family.

The fifth variant, NC_000017.11:g.44911435dup (NM_002055.5:c.928dup) within the coding region of *GFAP* is predicted to result in a frameshift at codon 310, and termination after incorporation of 112 aberrant amino acids (p.Met310Asnfs*113) (wild type GFAP is 431-438 aa). Furthermore, analysis of additional family members IV-6, V-8, VI-3, and VI-4 showed that the variant in *GFAP* segregated with the optico-retinal phenotype in the family, although one case of non-penetrance was observed (individual IV-6). The variant is not present in the 125,748 exomes and 76,156 genomes of Genome Aggregation Database (gnomAD v.2.1.1) and it is classified as “likely pathogenic” according to ACMG guidelines (PVS1_moderat, PM2_supportive, PP1_strong).

Together, our clinical and genetic analysis show that the GFAP variant p.Met310Asnfs*113 cause a novel human eye phenotype and suggest that GFAP might be involved in human retinal development.

### Expression of GFAP in retinal neural progenitors of the developing human eye

To determine if GFAP is involved in human retinal development, we performed immunohistochemical analysis of human embryonic and fetal tissue sections at different stages of eye development. We observed strong expression of GFAP in retinal neural progenitors (RNPs) as early as 35 days post conception, dpc (Fig. 2a-b).

**Figure 2.**
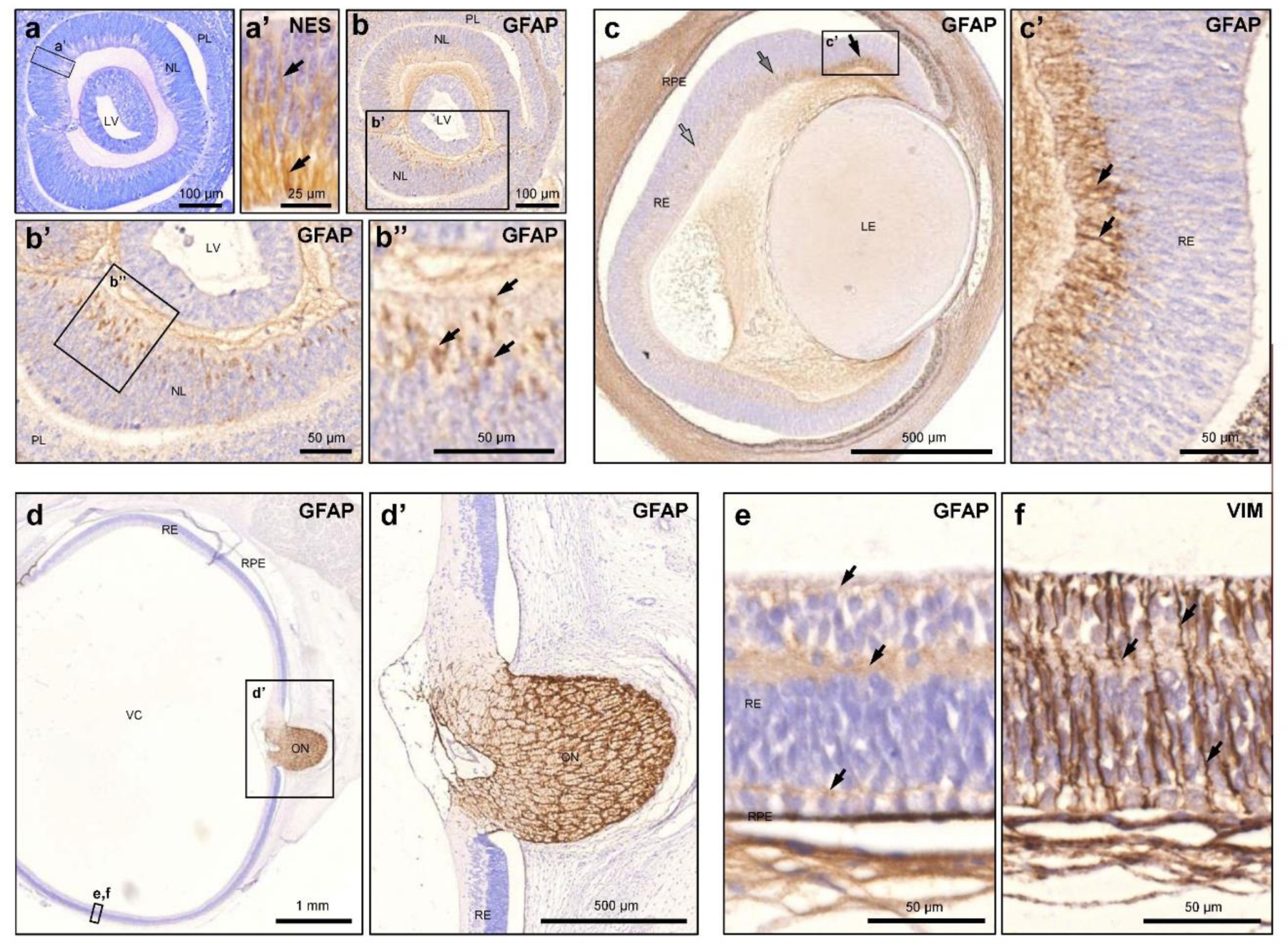
GFAP is expressed in human retinal neural progenitors. **a-b.** Sagittal section of human eye anlage at 35 dpc. **a.** Toluidine blue stained section. **a’.** Nestin (NES) immunostaining marks retinal neural progenitors (RNPs) in the NL. **b, b’, b’’.** Immunostaining shows strong expression of GFAP in RNPs within the NL. **c.** Sagittal section of human eye at 51 dpc. GFAP immunostaining shows strong expression in early RNPs (black arrow), and weaker expression in more differentiated RNPs (grey arrows). **d-f.** Sagittal section of human eye at 13 wpc. Immunostaining show very weak GFAP expression (**d, e,** arrows) in MGC, marked by Vimentin (VIM) expression (**f**, arrows), but very strong GFAP expression in astrocytes, entering the retina from the optic nerve. LV: lens vesicle, LE: lens, NL: nervous layer, ON: optic nerve, PL: pigment layer, RE: retina, RPE: retinal pigment epithelium, VC: vitreous cavity.

At 51 dpc we observed strong GFAP expression in early RNPs near the ciliary body and gradually weaker expression in more differentiated RNPs in regions closer to the optic nerve papilla (Fig. 2c). Immunostaining of a 13 weeks post conception eye showed very weak GFAP expression in Müller Glia Cells, but strong expression in astrocytes entering the retina from the optic nerve (Fig. 2d-f).

### The c.928dup variant does not result in extensive GFAP protein aggregation

A hallmark of AxD pathology is the presence of RFs, which are cytoplasmic protein aggregates in astrocytes of the brain, formed predominantly due to gain of function missense variants in the *GFAP* gene^12,13^. To assess potential protein aggregation properties of the p.Met310Asnfs*113 GFAP variant, we constructed plasmids, capable of expressing GFP-tagged human wild-type (WT) or p.Met310Asnfs*113 GFAP variant under the control of a zebrafish *gfap* promoter. The constructs were injected into fertilized zebrafish oocytes and the resulting number of GFAP protein aggregates within the brain and spinal cord was quantified^23^. For comparison, we injected a construct with a previously reported GFAP missense variant (p.Arg79Cys) associated with AxD and known to cause aggregation in the zebrafish *in vivo* assay^11,23^. As expected, the p.Arg79Cys GFAP variant resulted in extensive protein aggregation in the spinal cord and brain compared to the WT GFAP (Fig. 3a, 3a’). In contrast, we found that there was no significant difference in the number of aggregates between WT and p.Met310Asnfs*113 GFAP variant (Fig. 3a, 3a’). Western blot (WB) analysis confirmed the presence of p.Met310Asnfs*113 protein indicating that the absence of GFAP aggregates was not due to extensive protein degradation *in vivo* (Fig. 3b). These results suggest that the disorder observed in affected individuals with the c.928dup *GFAP* allele has a different pathological mechanism than that seen in AxD patients.

**Figure 3.**
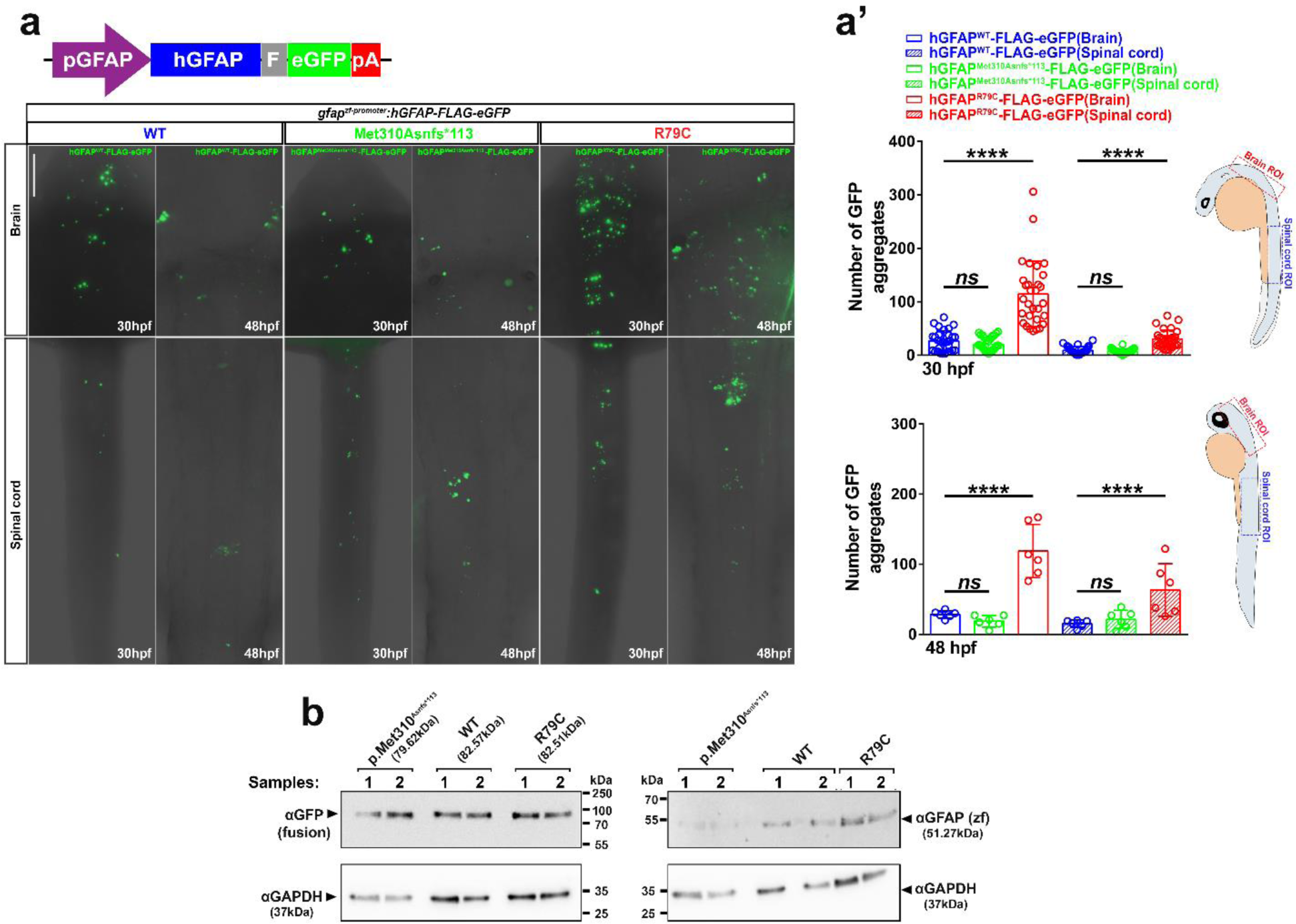
The p.Met310Asnfs*113 variant protein have similar aggregation properties as WT GFAP in vivo. **a.** Diagram of the injected expression plasmid encoding human GFAP C-terminally fused to a FLAG epitope and EGFP driven by a zebrafish *gfap* promoter. pGFAP: zebrafish gfap promoter; hGFAP: human GFAP; F: 3× FLAG epitope tag; eGFP: enhanced green fluorescent protein and pA: polyadenylation sequence (**a**, top). High-magnification dorsal view ROI images of the head and trunk for each injected larvae group, scale bar: 100 μm (**a**, bottom). **a’.** Quantification of GFAP aggregates in the brain and spinal cord of 30 hpf and 48 hpf larvae microinjected with 50 pg GFAP(WT), GFAP(p.Met310Asnfs*113) or GFAP(R79C) fusion construct (30 hpf: n=30 larvae for each genotype; 48 hpf: n=6 larvae for each genotype). **b.** Western blot analysis with GFP, zebrafish Gfap and GAPDH antibodies of the same injected groups as in (**a**); antibodies and protein size calculation as indicated. The data is presented as mean±sd. Asterisks indicate P-values obtained by using One-way ANOVA test: ****: P < 0.0001, ns: not significant.

### Silencing of *gfap* expression in zebrafish causes a retinal dysplasia-like phenotype

To investigate whether *GFAP* loss of function disrupts eye development *in vivo*, we used the CRISPR/Cas9 system to silence *gfap* expression and diminish potential genetic compensation^24^. We identified and characterised two phenotypically similar alleles with a 460 bp (*gfap^Δ^*^460^) and 560 bp deletion (*gfap^Δ5^*^60i^*^ns^*^24^), respectively, spanning the TATA-box among other elements in the *gfap* promoter region as well as the start-codon in the first exon (Fig. S1). These *gfap* promoter-less mutants showed an expected reduction in *gfap* mRNA and protein expression levels found in the optic nerve fibre layer, MGCs processes and endfeet (Fig. S2). The results outlined below are generated with the *gfap^Δ5^*^60i^*^ns^*^24^ mutant allele, but for the examined phenotypes in the *gfap^Δ4^*^60^ allele we observed similar phenotypes (Fig. S3).

The eyes of *gfap^+/-^* and *gfap^-/-^* larval showed a reduction in size at six days post fertilization (dpf), but not earlier, without affecting the body length (Fig. 4a-b’ and Fig. S4a-b’). Rescue experiments with human WT and c.928dup *GFAP* mRNA demonstrated that WT *GFAP* mRNA (*hGFAP*^WT^), but not c.928dup mRNA rescued the eye size phenotype (Fig. 4c, c’). Sagittal sections of the *gfap* mutant larvae showed normal cells layers in the retina, but at 6 dpf revealed a significant decrement in cells of the ganglion cell layer (GCL), inner nuclear layer (INL) and photoreceptor layer (Fig. 4d, e). Similarly, in adult fish the retina was smaller in *gfap* mutant fish due to thinner GCL, INL and outer segment (OS) (Fig. 4d, f). An increment in apoptosis in the inner layers of the eye and around the lens was observed in the mutants as early as 52 hours post fertilization (hpf), which could contribute to the reduction in eye size (Fig. 4g).

**Figure 4.**
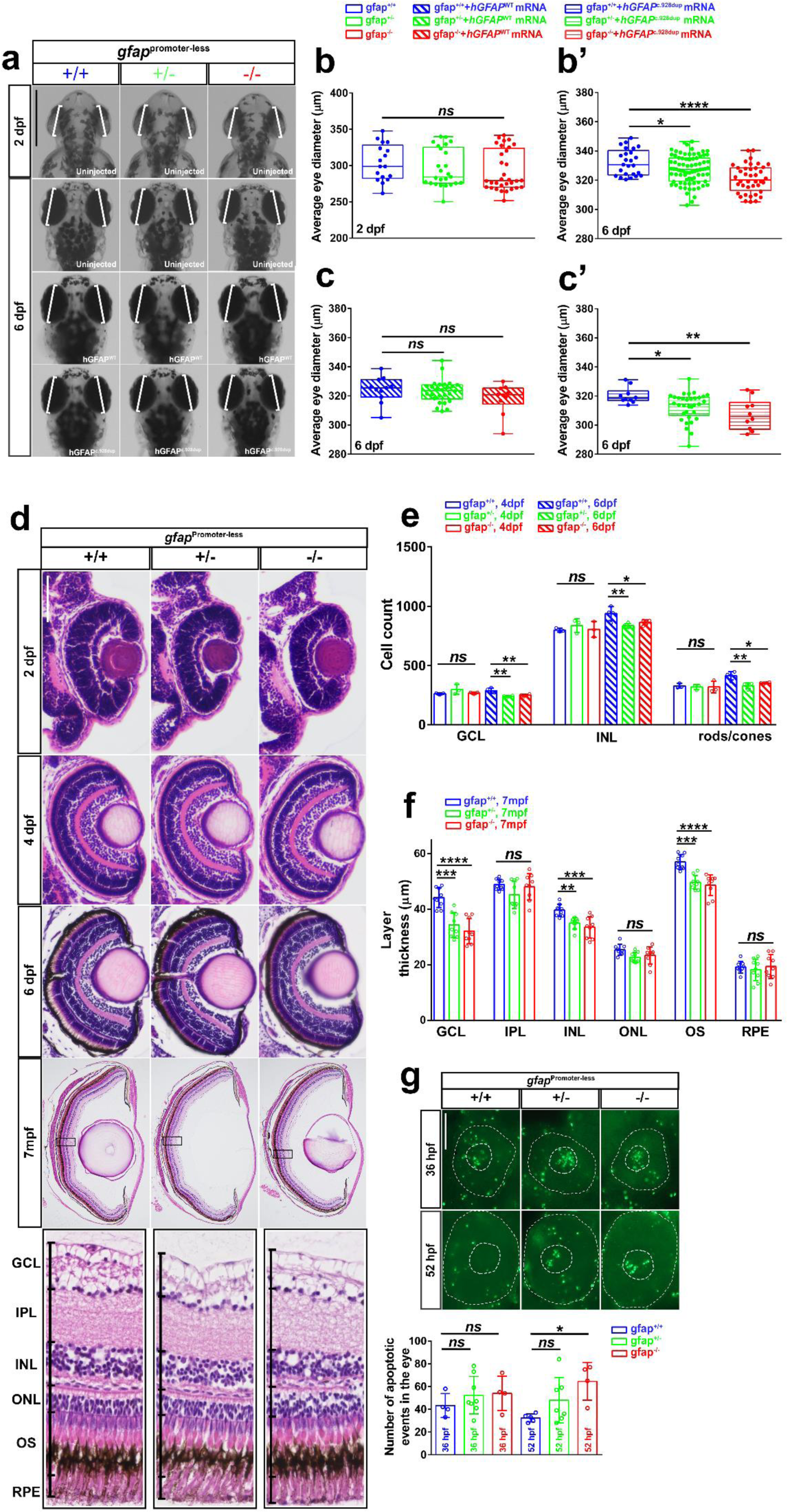
Zebrafish promoter-less *gfap* mutants have smaller eyes which can be rescued with WT but not c.928dup mRNA. **a.** Dorsal view images of 2 and 6 dpf larvae head of WT and *gfap* mutant siblings with or without injection of 100pg WT *gfap* mRNA or c.928dup mRNA (as indicated). White bars indicate an approximation of the major axis diameter for comparison. Scale bar: 50 μm. Measured mean diameter of both eyes for uninjected 2 dpf (**b**) 6 dpf (**b’**) larvae (n=17-32 and n=33-80 for each genotype, respectively) and 6 dpf larvae injected with WT (**c**) and c.928dup mRNA (**c’**) (n=10-26 and n=10-24 for each genotype, respectively). **d.** Hematoxylin and eosin staining of WT and *gfap* mutant siblings eyes at different stages: 2, 4 and 6 dpf and seven months post fertilization (mpf, adult fish), scalebar: 50 μm and scale bar: 500 μm, respectively (**d**, top). Higher magnification of retinal layers in 7 mpf fish. GCL, ganglion cell layer; IPL, inner plexiform layer; INL, inner nuclear layer; OPL, outer plexiform layer; ONL, outer nuclear layer; OS, outer segments; RPE, retinal pigment epithelium, scale bar: 50 μm. (**d**, bottom). **e.** Quantification of cells in the GCL, INL and photoreceptor layer from 4 and 6 dpf WT and *gfap* mutant sibling retina sections. **f.** Mean layer thickness in 7 mpf retina sections. **g.** TUNEL assay of 36 hpf and 52 hpf eyes in the same group of WT and *gfap* mutant siblings (n=4-8 and n=4-7 for each genotype, respectively), scale bar: 100 μm (**g**, top). Quantification of apoptotic events in the eye of the same group (**g**, bottom). The data are presented as mean±sd. Asterisks indicate P-values obtained by using One-way ANOVA test: *: P < 0.05, **: P < 0.01, ***: P < 0.001, ****: P < 0.0001, ns: not significant.

We hypothesized that the structural changes in eye morphology in mutants might cause vision impairment. We tested visual behaviour for zebrafish *gfap*^+/-^ and *gfap*^-/-^ mutants and wildtype siblings, by determining reflective behaviour to sudden changes in the light conditions (startle response) and reflective ability to position themselves stably in their environment (optomotor response (OMR)). The startle response protocol consisted of seven light to dark switches (Fig. 6a). During the testing period we observed that the distance travelled by *gfap*^+/-^ and *gfap*^-/-^ larvae was less than their WT siblings (Fig. 5a and Fig. S5) and, importantly, less than 75% on average of the mutant larvae exhibited a startle response as compared to over 90% observed in WT siblings (Fig. 5b). We observed similar indication of vision impairment when assessing the OMR of *gfap*^+/-^ and *gfap*^-/-^ larvae, which showed increased inactivity and decreased duration of high-velocity swimming, when compared to WT siblings (Fig. 5c). Together, the behaviour assays support that the vision of the mutant larvae is impaired.

**Figure 5.**
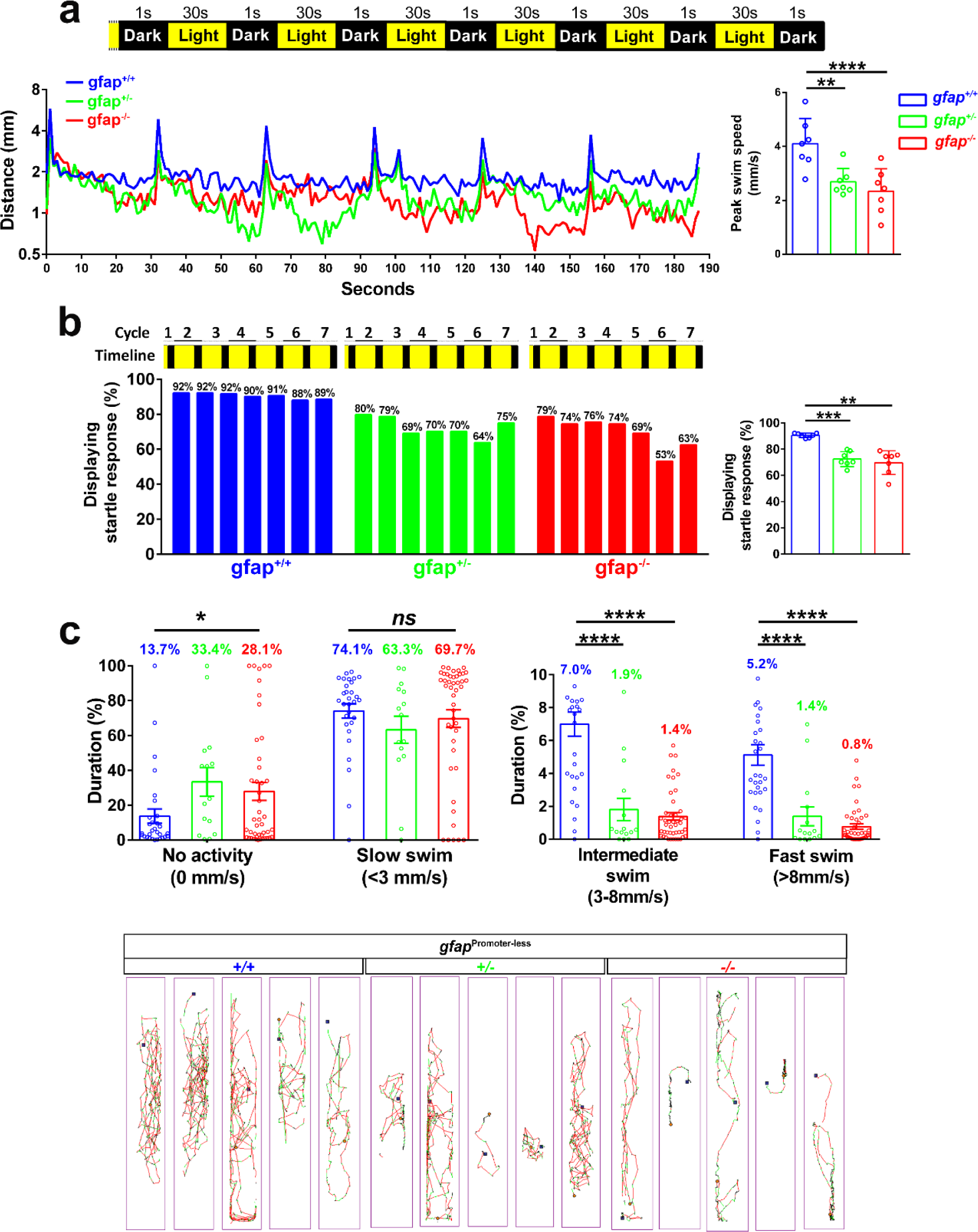
*Gfap* promoter-less mutants are visually impaired. **a.** Startle-response assay. Diagram of the 3 minutes and 7 seconds testing protocol following adaptation to light consisting of seven dark periods and six light each with a duration of 1 second and 30 second, respectively (**a**, top). Plot of the mean distance travelled by 7 dpf WT and mutant siblings over the testing phase (n=48-96) (**a**, bottom left). Bar plot of the mean maximum swim speed achieved during light switches (**a**, bottom right). **b.** Distribution of the mean larvae number that respond to light switches (**b**, left). One-way ANOVA test of the percentage responsive larvae during all the cycles (**b**, right). **c.** OMR. Bar plot of the duration spend at different swim speed: no activity, slow (<3mm/s), intermediate (3-8 mm/s) and fast (>8mm/s) (n=15-45) (**c**, top). Representative locomotion of five 9 dpf larvae color-coded for swim speed (slow: black, intermediate: green, fast: red) for WT and *gfap* mutant siblings (**c**, bottom). Data is presented as mean±sd. Asterisks indicate P-values obtained by using One-way ANOVA test: *: P < 0.05, ****: P < 0.0001, ns: not significant.

**Figure 6.**
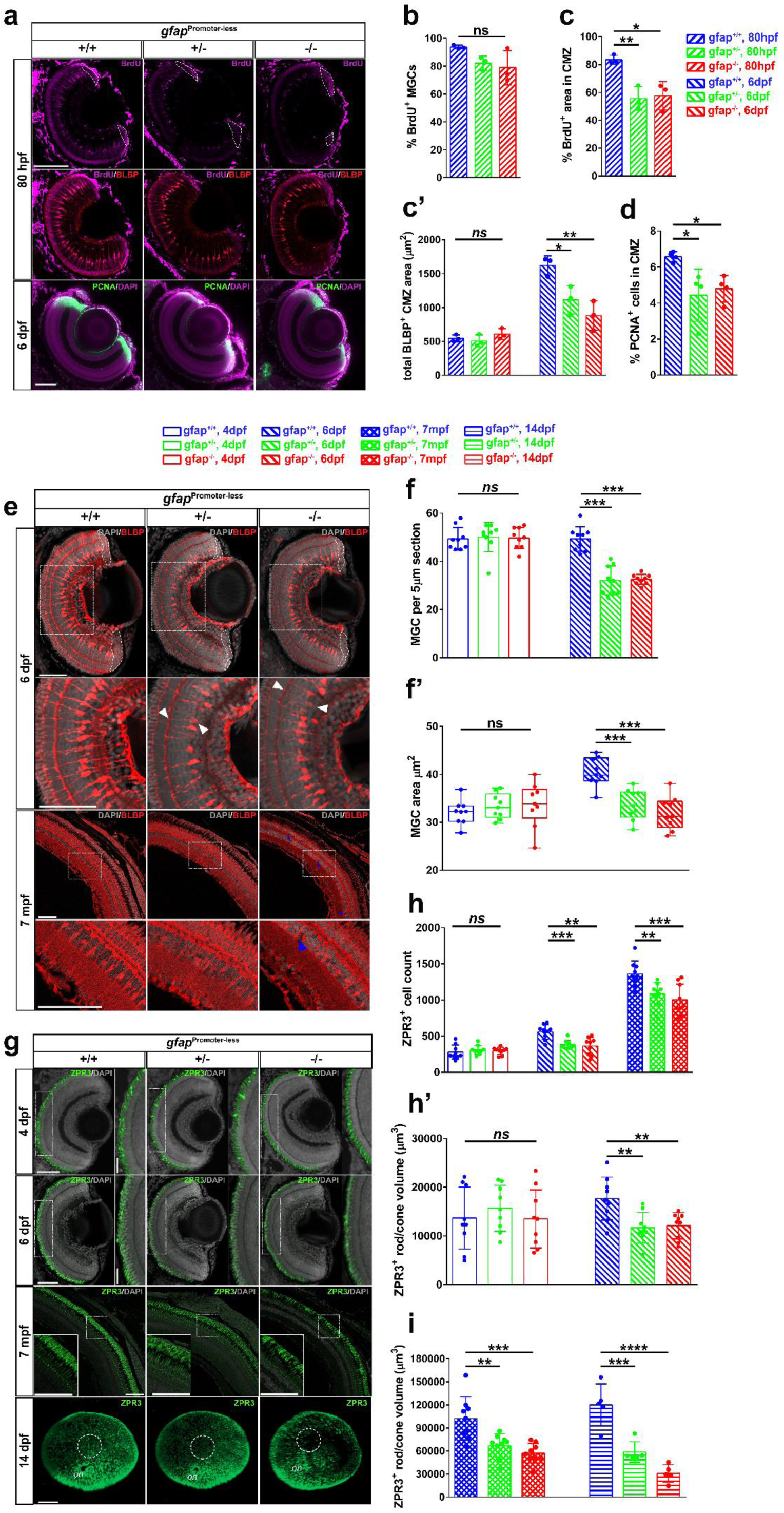
MGCs, rods and double cones are abnormal in *gfap* promoter-less mutants. The BLBP marker was used to label progenitors in the CMZ and MGCs^27,28^. **a.** IF maximum projection of BLBP^+^ MGCs and BrdU^+^ cells in the center of the eye from 80 hpf WT siblings and *gfap* mutants (**a**, top). White lines indicate the BLBP^+^ CMZ area. IF maximum projection of CMZ PCNA^+^ cells in the center of the eye from 6 dpf WT siblings and *gfap* mutants (**a**, bottom). Scalebar 50 μm. **b.** Bar plot of percent BLBP^+^ and BrdU^+^ MGCs in 80 hpf whole eyes from WT siblings and *gfap* mutant. **c.** Bar plot of percent BrdU^+^ area normalized to CMZ BLBP^+^ area. **c’.** Bar plot of mean CMZ BLBP^+^ area (dorsal and ventral) measured in sections from 80 hpf and 6 dpf WT siblings and *gfap* mutants. **d.** Bar plot of percent PCNA^+^ cells of total DAPI^+^ cells in the retina of 6 dpf WT siblings and *gfap* mutant retinal sections. **e.** IF maximum projection of BLBP^+^ MGCs in 6 dpf (**e**, top) and 7 mpf (**e**, bottom) WT and *gfap* mutant retina. White lines indicate the BLBP^+^ CMZ area in 6 dpf retina. Higher magnification of MGCs at INL in ROI (white boxes). White arrowheads indicate weak BLBP expression in MGC soma and apical processes at OS. Blue arrowheads indicate tearing with no DAPI staining in adult retina (**e**, bottom). **f-f’**. Bar plot of the MGC count and measurements of soma area of MGCs in 4 and 6 dpf retinas from the same group of fish. **g.** IF maximum projection of Zpr3^+^ rods and double cone outer segment of 4 dpf, 6 dpf and 7 mpf WT and mutant retina (**g**, top and middle). Higher magnification of Zpr3^+^ rods and double cone outer segment in ROI (white boxes). 3D reconstruction of juvenile (14 dpf) eyes stained for Zpr3^+^ rods and cones (**g**, bottom). White circle highlights the pituitary macular region of the eye. **h-h’.** Bar plots of Zpr3^+^ cell number and volume in 4 and 6 dpf sections and 7 mpf from the same group of fish. **i.** Bar plot of Zpr3^+^ cell volume in sections and whole eyes, respectively. Scalebars: 50 μm (80 hpf, 4 and 6 dpf). Scalebars: 100 μm (14 dpf and 7 mpf). Data analysis from sections and whole eyes is n=9-12 sections and n=3-5 eyes for each genotype, respectively. Data is presented as mean±sd. Asterisks in plots indicate P-values obtained by using One-way ANOVA test: **: P < 0.01, ***: P < 0.001, ****: P < 0.0001, ns: not significant.

GFAP expression was previously identified in RNPs at early developmental stages and in MGCs at later stages in embryos obtained from *S. canicula* (small-spotted catshark)^5^. We investigated if GFAP loss affects MGCs and RNPs during retinogenesis in zebrafish. To assess this, *gfap* mutants were BrdU-labelled from 55 hpf shortly after the first appearance of Gfap expressing MGCs to 80 hpf, after cell proliferation is restricted to the CMZ^25,26^. The BLBP marker was used to label both progenitors in the CMZ and MGCs^27,28^. During the time of BrdU labelling, the MGC formation was not significantly affected, however a reduction in BrdU labelled cell area at the CMZ was apparent without change in BLBP^+^ progenitor cell area (Fig. 6a-c’). In contrast, at 6 dpf, a reduction in BLBP^+^ progenitor cell area was observed concurrently with reduced PCNA^+^ cells at the CMZ (Fig. 6a, c’ and d). This suggests that GFAP is necessary for normal cell proliferation and maintenance of progenitor pool at the CMZ but not the initial MGC proliferation and formation after initial retinogenesis.

A visible reduction in BLBP^+^ MGCs at the INL and a change in morphology accompanied by a general reduction in BLBP fluorescence intensity levels at the ONL and OS was observed in the *gfap*^+/-^ and *gfap*^-/-^ mutants both in larvae and adult fish (Fig. 6e, f, f’ and Fig. S4c, d). In mutant larvae, the MGCs that remained in the INL had reduced cell body area, thin processes and fewer apical branches at the base of rods and cones in the ONL/OS and adult sections showed evidence of tearing (Fig. 6e). Together this suggests that MGCs in the *gfap* mutants become dysfunctional during postembryonic growth of the eye.

Progenitors in the CMZ and MGCs are responsible for generation and maintenance of rod, cone and other neuronal lineages in the fish eye after initial retinogenesis is completed, while MGCs in the mammalian eye have a role in retinal homeostasis^8,26,29^. Given the observed MGC phenotypes in *gfap*^+/-^ and *gfap*^-/-^ mutants, we examined rod and double cone development during early postembryonic development and in the adult eye^30^.

We observed a significant reduction in Zpr3^+^ rods and double cones in *gfap* mutants at 6 dpf and in juvenile and adult fish (Fig. 6g-i and Fig. S4c’, d’, e), but not earlier. Although non-primates like zebrafish do not have a macular region, they have regions with high photoreceptor density similar to the human macular^31^. To determine whether Zpr3^+^ photoreceptor loss occurred at a pituitary macular region, we estimated the region relative to the optic nerve in our *gfap* mutants after performing 3D reconstruction of whole juvenile (14 dpf) and adult eyes (9 mpf). In *gfap* mutant juvenile eyes, visibly fewer photoreceptors in the pituitary macular region were observed than around the optic nerve and periphery regions (compared to WT siblings) (Fig. 6g). In contrast, Zpr3^+^ photoreceptor cell loss was observed both in the periphery regions and the pituitary macular region of adult *gfap*^+/-^ and *gfap*^-/-^ eyes, with *gfap*^-/-^ being more affected than *gfap*^+/-^ (Fig. S6).

In summary, clinical and genetic analyses of patients revealed a rare GFAP variant associated with a novel human eye phenotype. Expression analyses and *in vivo* experiments supported a previously unknown role of GFAP in retinal development.

## DISCUSSION

### Identification of a novel inherited optico-retinal dysplasia

The current study reports on a six-generation family with a unique hereditary optic nerve head dysplasia that was examined in six individuals from the last three generations. The eye disorder was characterized by intra- and pre-papillary glia-like tissue extending along the retinal vessels and involving the central retina with edema and atrophy of the retinal pigment epithelium (RPE) of the macula. OCT demonstrated extensive optic nerve head gliosis with accumulation of intra-papillary amorphous substance and fluorescein angiography (not shown) showed leakage from the capillary system in the macular area.

Pre-papillary and peri-papillary glia-like structures representing a variety of developmental anomalies are common findings in clinical ophthalmology. Among these persistent hyperplastic primary vitreous (PHPV)^32,33^ and Bergmeister papilla^34^ are well known clinical entities. The majority of developmental prepapillary and preretinal tissue formations is unilateral and sporadic, however may also be found in hereditary systemic disorders as neurofibromatosis type 2^35^ and familial exudative vitreoretinopathy^36^. PHPV and Bergmeister papilla are considered the result of a deficient regression of the primary hyaloid artery system, however, hypovascular glial overgrowth from the optic nerve head in foetuses indicates that non-vascular mechanisms also exist^37^. Combined hamartoma of the retina and retinal pigment epithelium^38,39^ shows similarity to the optico-retinal disorder described here, which however, had no sign of a hamartoma of the macular pigment epithelium. Similar clinical findings were described in a family showing autosomal dominant inheritance in three generations reported at the ARVO Annual Meeting in 2010^40^. Clinical examination of six affected family members from that family showed variable phenotypes with different degrees of fibro-glial proliferation over the optic discs and peripapillary area and various degrees and retinal pigment epithelium disruption and migration. A genetic analysis for causative variants in *NF2* was negative.

The described optico-retinal dysplasia is to our best knowledge the first eye disorder without systemic involvement associated with a variant in the glial fibrillary acidic protein (GFAP), a classical marker of glial cells, among which astrocytes have crucial functions in retinal development.

### Human GFAP expression pattern in RNP and MGCs suggest an evolutionary conserved role of GFAP in retinal development

In our study we examined GFAP expression during early human retinal development. Previous studies indicated that GFAP is expressed during human fetal retina development from 8 wpc but similar to the postnatal development in mice its expression was observed only in astrocyte precursors that had migrated to the optic nerve head^4,41–44^. In contrast, our findings demonstrate that robust GFAP expression is present in RNPs throughout the retina of the developing human fetal eye already at 35 dpc. Furthermore, our findings suggest that GFAP expression intensifies during astrocyte precursor maturation and migration into the vascularized central retina beginning at 13 wpc which is earlier than previously estimated (16-18 wpc)^42,45^. Interestingly at 51 dpc we observed strong GFAP expression at the periphery of the developing neural retina near the ciliary body. The gradual weakening of GFAP expression in RNPs with radial orientation and changes in cell morphology with RNPs differentiation is characteristic of the CMZ. The ability of CMZ to generate retinal cells during development has been thought to be lost in mammals^26,46–48^. However, a recent study in mice demonstrated that Cyclin D2^+^ and Msx1^+^ cells from the ventral embryonic CMZ can become retinal ganglion cells^49^ and all seven cell types in the eye^50^, respectively. Our results suggest that a subgroup of RNPs in the developing human eye matures into MGCs resulting in significant downregulation of GFAP expression with retained expression at distal processes and end-feet. The observed appearance of mature MGCs at 13 wpc is in agreement with a recent report dating it to 12 wpc^51^. To the best of our knowledge, a functional CMZ and GFAP expression in RNPs have not previously been demonstrated in primates, but appear to resemble the GFAP immunoreactivity pattern observed in the CMZ of embryonic catshark fish (*S. canicula*). In catshark fish, Gfap expression occurs in the pre-neurogenic retina in neural stem cells, which during the neurogenic developmental stage elongate and transform into MGCs when RNP change to an asymmetric mode of cell division^5^.

### GFAP c.928dup does not cause extensive protein aggregation in vivo

Genetic analysis of the affected individuals demonstrated that the disorder segregates with a variant in *GFAP* (c.928dup; p.Met310Asnfs*113) causing a frameshift and incorporation of 112 aberrant amino acids resulting in a partial deletion of the 2B rod domain. The observed dysplasia was stationary across decades in the examined individuals. Affected individuals were in good health, with none of the classical indications of severe AxD with the exception of mild difficulties in swallowing, which can be found in the clinical spectrum of AxD^13^.

AxD is a heterogeneous disease that can be mild or even asymptomatic despite remarkable atrophy of the brainstem and spinal cord on MRI^52^. In fact, a nonsense mutation variant in proximity of our variant has been reported to cause a mild and atypical case of AxD (c.1000G>T, p.E312*)^14,53^. Overexpression of GFAP without the tail domain has previously been reported to form polymorphic aggregates^54,55^. However, the GFAP variant p.Met310Asnfs*113 has a large number of out-of-frame amino acids, and thus the outcome of this variant is unpredictable. Furthermore, GFAP aggregation and AxD outcome is complicated by the expression of different GFAP isoforms and mis-splicing in humans, which are difficult to model in animals^56,57^. Several isoforms of GFAP exist in humans, however, not all of them are translated into protein product^58^. Two isoforms (Δexon6 and Δ135) are expected not to be affected by the c.928dup variant, and consequently may contribute to the mild clinical signs in the affected individuals. Lastly, at present we cannot rule out that the c.928dup *GFAP* variant produces transcripts that undergo nonsense-mediated-decay (NMD) in the affected individuals^59^, but since the novel stop-codon is only 31 nucleotides from the wild type GFAP stop-codon (and in the same exon) we find NMD unlikely. Taken together, we cannot exclude the possibility that the affected individuals with the optico-retinal dysplasia represent asymptomatic cases of AxD. However, given that the p.Met310Asnfs*113 variant does not result in extensive GFAP aggregation and the disease progression is largely stationary in the affected individuals, we find this scenario unlikely.

### Retinal dysplasia-like phenotype in zebrafish gfap mutants

To examine the consequence of gfap loss-of-function, we generated a promoter-less mutant to abolish potential genetic compensation^24^. Several studies of *GFAP* null mice reported normal development, growth, fertility and lifespan^60–64^. Although obvious candidates for compensation (such as Vimentin and Nestin) were ruled out^61,65,66^, recent studies in vertebrates have proven genetic compensation to be more complex, for instance as the result of degradation of mutant mRNA and upregulation of genes with sequence homology to mutant mRNA^24,67^. Characterisation of our zebrafish promoter-less *gfap* mutants revealed a previously uncharacterized small-eye phenotype and visual impairment at post-embryonic stages. Cell proliferation changes at the germinal zone located all around the retinal margin and cell shrinkage may influence neurogenesis and retina size in the larvae and adult fish^68^. Our data suggest reduced cell proliferation at the CMZ is the main contributor to the phenotype in *gfap* mutants. MGCs in the fish eye proliferate at low rates to produce neural progenitors during postembryonic development of fish retina^29^ and provide tensile strength^69^, suggesting that the MGC phenotype in *gfap* mutants could contribute to the observed small-eye phenotype and photoreceptor loss. However, the initial MGC formation was not affected in the mutants probably due to the intact BLBP^+^ progenitor pool in the CMZ. In contrast, later in development MGC phenotypes were evident concurrently with a depletion in the progenitor pool at the CMZ. This indicates that loss of MGCs is a secondary effect to CMZ dysfunction occurring after initial retinogenesis is completed. These MGCs displayed smaller soma with abnormal processes and branching at the ONL, with weaker BLBP fluorescence intensity. In the brain, *Blbp* transcription is induced in radial glia by migrating neurons through Notch signalling and blocking BLBP function in radial glia causes a reduction in the number of radial processes extended^27,40^. Abnormally compressed, thin and dysfunctional MGCs, have previously been observed in stressed retina of Gfap and Vimentin knockout mice^70^.

Concurrently with abnormal BLBP marker levels in MGCs, photoreceptor outer segment loss as well as thinning of retinal layers in aging fish. Photoreceptor apoptosis and changes in retinal layer thickness and intraretinal neovascularization has previously been observed in studies utilizing conditional ablation techniques to ablate MGCs in adult mice^71,72^. Abnormalities in the inner retinal layer and the foveal containing high density of photoreceptors were similarly observed in the majority of OCTs from the examined affected individuals. However, the zebrafish *gfap* mutants did not show any sign of a vascularized epiretinal glial mass at the optic nerve and no salient deficiency was observed in BLBP^+^ retinal astrocytes at the GCL and nerve fibre layer. The absence of developmental defects at the optic nerve may be attributed to the fact that unlike mammals, zebrafish astrocytes of the optic nerve and many other fish species express Gfap transiently or not at all^73,74^. Furthermore, the progressive nature of the phenotype observed in the *gfap* mutant fish may be attributed to the fact that retinas of postembryonic teleost fish develop faster and exhibit continuous growth in the retina over their entire lifetime, while the human eye is morphologically fully developed by 2-3 years of life^28,75,76^.

In conclusion, this work identifies a GFAP mutation causing optico-retinal dysplasia and vision impairment likely through haploinsufficiency or a dominant negative mechanism. We further demonstrate that RNPs during human retinogenesis express GFAP and that uncompensated loss of GFAP in zebrafish results in detrimental changes for MGCs and photoreceptors. The role for GFAP as a contributor to early retinogenesis calls for care when designing clinical studies and therapeutic approaches aiming to abate GFAP function in AxD patients.

## MATERIAL AND METHODS

### Antibodies and PCR primers table

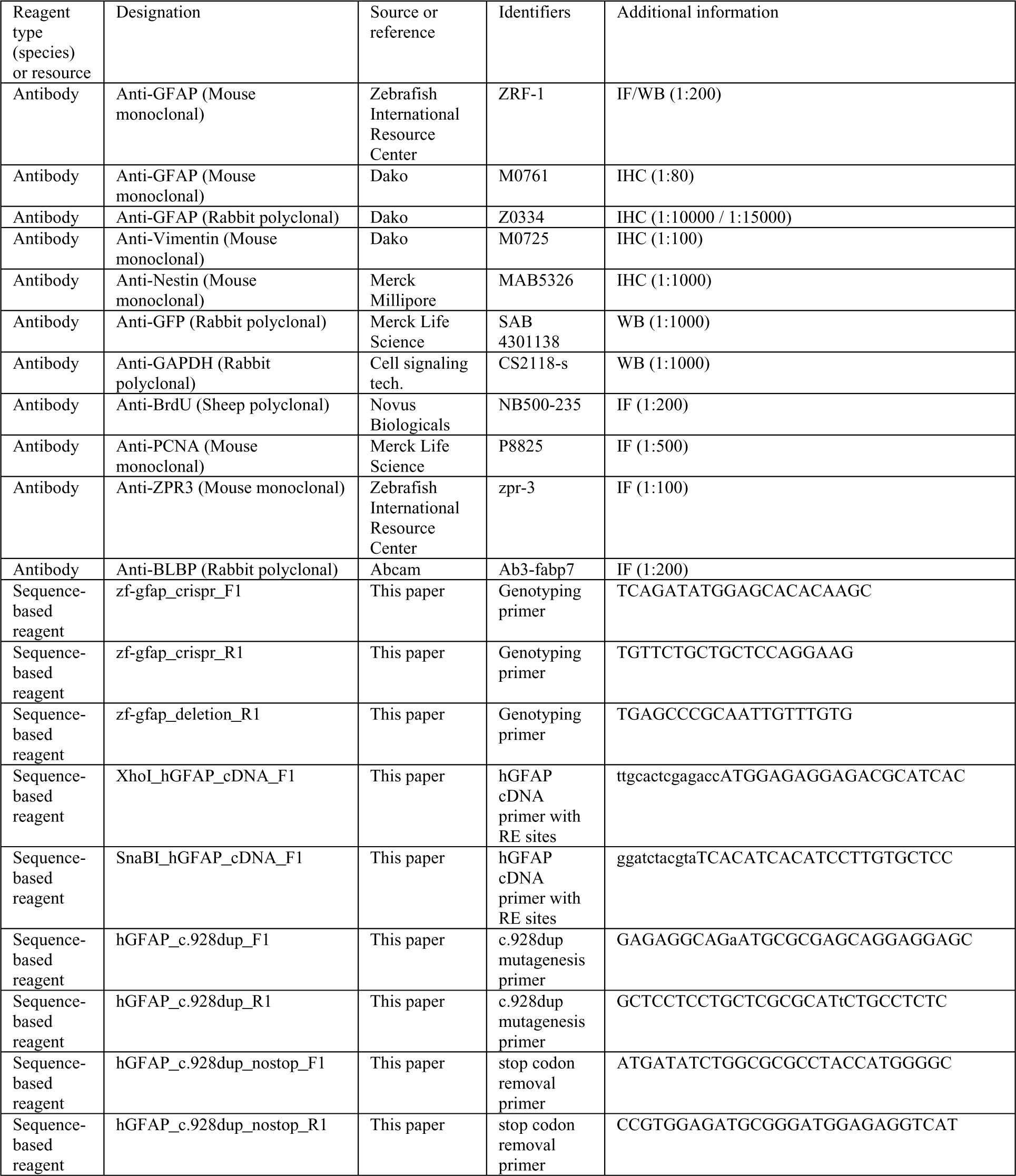

### Family pedigree and clinical investigations

The pedigree encompassed six generations, the first three individuals were known to be visually impaired by history only, three were followed for a period of 35 years. All available patient files were retrieved and scrutinized and a re-investigation of five affected, one unaffected, and one non-penetrant individual was made. Re-examinations took place at the Department of Ophthalmology, Rigshospitalet, Copenhagen.

Standard ophthalmological examination was performed in all seven individuals including best corrected visual acuity (BCVA) according to the ETDRS procedure. The subjects underwent fundus photography (Topcon), spectral-domain-optic coherence tomography (SD-OCT) (Heidelberg Eye Explorer) including autofluorescence photography, one of the subjects had fluorescence angiography, indocyanine green angiography, adaptive optics, and full-field ERG.

Written informed consent was obtained from all participants. The study was approved by the National Committee on Health Research Ethics, Denmark (journal number 1301394) and followed the tenets of the Declaration of Helsinki.

### Molecular genetic analysis

Blood samples were obtained for eight family members (IV-4, IV-6, V-1, V-3, V-7, V-8, VI-3, VI-4) and high molecular DNA was extracted using the Chemagic 360 machine (Perkin Elmer, Waltham, Massachusetts, USA). Whole genome sequencing was performed by BGI Group (Shenzhen, China) using Illumina HiSeq X-ten. A mean coverage of 30 X were provided. Alignment was performed to NCBI hg19 version of the human genome using BWA software. GATK (Broad Institute, MIT Harvard, Cambridge, MA, USA) was used for variant calling (SNV and indels). Three affected (V-1, V-3, V-7) and one unaffected (VI-3) individual were analyzed. A total of 398,394 unique variants were retrieved for the four samples. The VarSeq program (Golden Helix, Bozeman, Montana, USA) was used for filtering. Coding variants (including 8 bp flanking intron sequence) with a minor allele frequency (MAF) below 0.5% in the gnomAD v2.1.1 database (https://gnomad.broadinstitute.org/) and an allelic frequency between 0.3 and 0.7 were retrieved. The zygosity of affected individuals were set to heterozygous whereas the unaffected individual was set to reference. The Alamut Visual Plus program (Lausanne, Switzerland and Boston, Massachusetts, USA) were used for further analysis of variants.

Segregation analysis was performed using primers GFAP-ex6-FH (5’-acccactgcttactggcttatcGAGATGCCAGGGGAGAAGG-3’) and GFAP-ex6-RH (5’-gaggggcaaacaacagatggcCAGGGCCAGCTTGACATTG-3’) for PCR amplification and subsequent Sanger sequencing. Capital letters are gene specific and small letters are used for subsequent sequencing. Sequencing was performed using BigDye v3.2 terminators (Applied Biosystems, Waltham, Massachusetts, USA) and analysis on an ABI3130XL (Applied Biosystems).

Details of the primary antibodies including dilutions and suppliers are listed in Antibodies and PCR primers table. Control sections were incubated with mouse IgG1 or irrelevant rabbit antibodies, as well as subjected to omission of primary or secondary antibodies. These were always blank.

### Immunohistochemistry of human embryonic and fetal eyes

Four human embryos and 5 fetuses were obtained from legal abortions after informed consent from all contributing women following oral and written information, in accordance with the Helsinki Declaration II, and was approved by the Danish Regional Committee on Health Research Ethics (KF–V.100.1735/90) & (KF-11 2006 – 4838).

Immediately following the abortion, entire embryos of 35 dpc, 51 dpc and 53 dpc (days post conception) and fetal eyes from fetuses of 13, 15 and 19 wpc (weeks post conception) were removed and promptly fixed for 12–24 h at 4 °C in either 10% neutral buffered formalin, 4% formol-calcium, Lillie’s or Bouin’s fixatives. This procedure kept the time from delivery to fixation at a minimum, normally less than 2 h, in order to retain optimal tissue quality. Prior to paraffin embedding, the specimens were dehydrated with graded alcohols, and cleared in xylene. Then 3 - 6 μm thick serial sections were cut in transverse, sagittal or horizontal planes, and placed on silanized glass slides.

Sections were deparaffinized and rehydrated in xylene following standard protocols. Endogenous peroxidase was quenched using a 0.5% solution of hydrogen peroxide in TBS for 15 minutes. Following rinses with TRIS buffered saline (TBS, 5 mM Tris-HCl, 146 mM NaCl, pH 7.6), non-specific binding was inhibited by incubation for 30 minutes with 10% goat serum at room temperature. The sections were incubated overnight at 4°C with primary antibodies (Antibodies and PCR primers table) diluted in blocking buffer and washed with TBS. For bright field light microscopy analysis, the REAL EnVision Detection System (Peroxidase/DAB+ rabbit/mouse, code K5007, DakoCytomation, Glostrup, Denmark) was used for detecting mouse and rabbit primary antibodies. The sections were washed with TBS. Positive staining was recognized as a brown color. The sections were counterstained with toluidine blue or Mayers hematoxylin and dehydrated in graded alcohols and coverslipped with Pertex mounting media.

### Zebrafish husbandry

The AB Wild-type (WT) zebrafish strain was obtained from the Zebrafish International Resource Center (ZIRC) and maintained in the animal facility of the University of Copenhagen. The WT were raised at 28°C and kept in a constant LD cycle according to standard protocols. All experiments and animal handling procedures were approved and conducted under licenses from the Danish Animal Experiments Inspectorate (Protocol code: P18-120/P20-387).

### Generation of promoter-less *gfap* mutants

The promoter motifs of the zebrafish *gfap* promoter were mapped using the Eukaryotic promoter database (https://epd.epfl.ch/). Based on the location of the motifs, crRNAs were designed and evaluated using the IDT CRISPR design (Integrated DNA Technologies) tool. A pair of crRNAs was selected with a target region upstream of the 5’ UTR (IDT Design ID: CD.Cas9.JPYK4256.AC) and downstream of the *gfap* start-codon, respectively (IDT Design ID: Dr.Cas9.GFAP.1.AL). The CRISPR-Cas9 ribonucleoprotein complex was assembled according to the manufacturer’s protocol for zebrafish embryo microinjections. Briefly, equimolar amounts of crRNAs and tracrRNA were mixed in a nuclease-free duplex buffer to create a 3 μM gRNA complex solution. The gRNA solution complex was mixed with an equal volume of 0.5 μg/μL HiFi Cas9 Nuclease V3 (IDT) protein solution diluted in Cas9 working buffer (20 mM HEPES-KOH, 150 mM KCl, pH 7.5). The gRNA/Cas9 mixture was incubated at 37°C for 10 min to assemble the ribonucleoprotein complex. Microinjections were performed using a pressurized injection system (Intracel, Picospritzer III) with needles pulled from filamented capillaries (WPI) on a Sutter Instrument P30-867 guided by an Intracel M-152 micromanipulator. Injection droplets were calibrated to ∼125 µm diameter at the start of injection, resulting in approx. 1 nl injection mix delivered into the cytoplasm of one-cell staged embryos. Experiments were done using F2 and subsequent generations. Primers used for genotyping and sequencing are listed in Antibodies and PCR primers table.

### Eye diameter measurements

Measurements of the left and right eye width were conducted blindly from Z-series of mRNA injected (100 pg) and uninjected heterozygous in-crosses from different pairs at 2 and 6 dpf. The Z-series were collected with a Zeiss Axio Zoom.V16 microscope equipped with a AxioCam 705 color camera (Interval thickness: 5.0 μm) from individual larvae anesthetized in 0.02% tricaine methanesulfonate (Sigma-Aldrich, A5040) and mounted in 3 % methylcellulose.

### Total RNA isolation and qRT-PCR

To determine the degree of endogenous *gfap* promoter silencing in the promoter-less mutants, RNA was extracted from a pool of homozygous outcrosses, and in-crosses of wildtype sibling and homozygous from three different pairs using the RNA isolation kit (Zymo Research). One microgam of total RNA was reverse transcribed using SuperScript™ II (Invitrogen, Carlsbad, CA), and oligo-dT primers (Invitrogen). To detect *gfap* cDNA (NM_131373.2), qRT-PCR was performed using Brilliant III Ultra-Fast SYBR® Green qPCR master mix (Agilent Technologies) with the primers 5′-GGATGCAGCCAATCGTAAT-3′ (forward) and 5′-TTCCAGGTCACAGGTCAG-3′ (reverse). The qRT-PCR reactions were performed in triplicate and were analyzed on an ABI 7500 fast system. The data was normalized against the average expression value of the housekeeping gene β-actin2.

### Western Blot

Prior to protein extraction, microinjected larvae (30 hpf) were manually dechorinated, anesthetized and deyolked in Deyolking buffer (55mM NaCl, 1.8mM KCl, 1.25mM NaHCO_3_) at 1100 rpm. Deyolked larvae were collected at 3000rpm, 4°C and rinsed several times in Wash buffer (110mM NaCl, 3.5mM KCl, 10mM Tris pH 8.5, 2.7mM CaCl_2_). The larvae were subsequently lysed in Protein extraction buffer (1% SDS and 10mM Tris pH 7.4) with proteinase inhibitors and flash-freezed. Western blot was carried out as previously described in Schrøder et al., 2011 using horseradish peroxidase-conjugated secondary antibodies. Blots were developed with FUSION-Fx chemiluminescence system (Vilber Lourmat) while images were processed and analysed in Photoshop CS6 and ImageJ (Fiji), respectively. Primary antibodies for western blot and immunofluorescence are listed in Antibodies and PCR primers table.

### Cloning of GFAP fusion and transcription of mRNA from full-length constructs

The GFAP p.Met310Asnfs*113 fusion construct was cloned by site-directed mutagenesis of the mini-Tol2 vector containing the human WT GFAP construct1 introducing the nucleotide duplication at position 928. Subsequently another round of site-directed mutagenesis was performed to remove the stop-codon generated by the frameshift and the remaining ORF. For mRNA generation the GFAP cDNA was amplified from a universal human cDNA library (Takara Bio) and subcloned into the pCS2+ vector using the XhoI/SnaBI sites. The c.928dup mutation was introduced by site-directed mutagenesis with the full-length vector as previously described for the fusion construct. Capped mRNA was generated *in vitro* after NotI linearization with the mMESSAGE mMACHINE Kit (ThermoFisher) kit and purified according to manufactures instructions. All plasmids were verified by DNA sequencing (ABI3130xl Genetic Analysers). Primers used for cloning are listed in Antibodies and PCR primers table.

### GFAP aggregation assay

The GFAP aggregation assay was performed as previously described1. Briefly, expression vectors containing the WT, p.Arg79Cys and p.Met310Asnfs*113 GFAP construct were microinjected into the cytoplasm of one-cell staged zebrafish embryos (50 pg). At 30 hpf the embryos were anesthetized and mounted in 3% methylcellulose for imagining. Z-series of the head and trunk regions were collected (Interval thickness: 4.0 μm) with a Zeiss Axio Zoom.V16 microscope equipped with a AxioCam 705 color camera. GFAP aggregates were blindly counted using the Cell Counter plugin in ImageJ (Fiji). Representative stacking images were assembled in ZEN 3.2 Pro (Zeiss) and Photoshop CS6 software (Adobe).

### Sectioning, H&E and immunofluorescence of the fish eye

For sectioning, 2, 4 and 6 dpf zebrafish larvae from heterozygous in-crosses were collected and fixed in 4% paraformaldehyde (PFA) overnight at 4 °C. Larvae were rinsed in PBS, genotyped and their heads embedded and orientated in 5% agarose. Genotyped adult fish were fixed in 4% PFA for several days at 4°C after anesthetizing and euthanizing the fish. The eyes were carefully enucleated, embedded and orientated in 5% agarose. The agarose blocks were dehydrated in increasing alcohol solutions over several days (70%/96%/99% EtOH). Following dehydration of the agarose block, the block was transferred into Xylene and into paraffin overnight at 60 °C. The block was embedded into fresh paraffin prior to sectioning and slide collection (Sagittal sectioning, thickness: 5 or 7 μm). Slides with the sections were dried overnight at 37 °C followed by standard haematoxylin and eosin or immunofluorescence (IF) staining using secondary antibodies coupled with Alexa488 (Invitrogen, 1:100) or RedX (1:100, Jackson ImmunoResearch) and nuclear stained with DAPI. Haematoxylin and eosin-stained slides were imaged on AxioScan 7 slide scanner (Zeiss) and IF-stained slides were imaged on LSM710 confocal system (Zeiss).

### TUNEL assay

For the TUNEL assay, embryos were fixed in 4% paraformaldehyde for 2 hours at room temperature and kept in 100% methanol overnight at -20°C. After gradual rehydration and genotyping, apoptotic cells were detected using the ApopTag peroxidase in situ apoptosis detection kit (Millipore). The samples were mounted in glycerol and Z-series of the right eye was collected (interval thickness: 3.0 μm) with Axio Zoom.V16 microscope equipped with a AxioCam 705 color camera (Zeiss).

### BrdU incorporation assay

BrdU injections were performed as done previously^8^. Briefly, 55 hpf larvae were lightly anaesthetized in E3 medium (0.05% MS-222; Sigma) and immobilized on an agarose plate. The larvae were subsequently microinjected into the middle of the yolk sac with 1.5 nL of 10mM BrdU in 15% DMSO/sterile saline solution. Injected embryos were returned to E3 medium and grown at 28°C until the time of fixation (80 hpf) and subsequently processed for ECi-DEEP tissue clearing.

### ECi-DEEP-Clear of the zebrafish eye

The clearing protocol for the eye was adapted from the DEEP-Clear protocol^77^. Unless otherwise stated the pH for solutions were neutral (pH 7-7.4). Zebrafish samples were first anesthetized with 0.04% tricaine methanesulfonate (Sigma-Aldrich, A5040) and euthanized. This was followed by immediate fixation in 4% paraformaldehyde at 4°C for 1-3 days and removal of fixative by rinsing in PBS several times at room temperature. For adult fish, eyes were carefully enucleated and dehydrated in pre-chilled acetone overnight at -20°C. Juvenile (and younger) fish were decapitated post-fixation and the whole head processed. After acetone treatment, samples were pre-treated with 3% H_2_O_2_ in Acetone for 30min at room temperature and gradually rehydrated in 75%, 50% and 0% acetone in PBS. In juvenile and adult eye samples, guanine crystals in the eye were dissolved by long rinses in PBS pH 6.0^78^. After several short dH_2_O rinses, samples were first depigmented by bleaching in 3% H_2_O_2_/1% KOH for approx. 45 min to 4 hours under strong light, followed by hydrophilic depigmentation in 8% THEED (Sigma-Aldrich, 87600) and 5% Triton-X in PBS pH 9-10, at 37°C for 30 min to 2 days. Depigmentation durations were dependent on the developmental stage of the eye. After several PBS rinses, unspecific antibody binding was blocked in PBDT (1% DMSO, 1% BSA, 0.1% Tween20, 0.5% Triton X-100, 5% goat serum, PBS) and samples incubated with primary antibodies in PBDT overnight at 4°C (Antibodies and PCR primers table). Unbounded antibodies were subsequently removed by several PBSTT (0.1% Triton-X, 0.1% Tween20, PBS) rinses at room temperature and incubated with secondary antibodies coupled to Alexa488, Alexa647 (Invitrogen, 1:100) or RedX (1:100, Jackson ImmunoResearch) in PBDT overnight at 4°C. Following several PBSTT rinses, the samples were nuclear stained with SYTOX Deep Red Nucleic Acid Stain (Thermo Scientific, S11381) or DAPI at room temperature and mounted in 2% low-melting agarose. Samples were then gradually dehydrated in 20%, 40%, 60%, 80%, 96% and 99% ethanol in dH_2_O (pH 9-10) at room temperature and immediately RI matched in ethyl cinnamate (Sigma-Aldrich, 8.00238) prior to imaging. Larvae and juvenile samples were imaged on a Zeiss CellObserver Spinning Disk confocal system using a Plan-Apochromat 20x/0.8 M27 objective or Zeiss Lsm900 using a Plan-Apochromat 40x/1.4 objective. Adult samples were imaged on Zeiss Light Sheet 7 using a Plan Apo 10x/0.5 objective. Deconvolution and correction of chromatic aberrations (if any) was done using Huygens Professional software (Scientific Volume Imaging).

### Startle response and optomotor response (OMR) assay

Prior to conducting the assay, fed larvae were cultured in a light environment for at least 1 hour. For startle response assessment, 7 dpf larvae were carefully transferred to a 48-well plate with 1 mL E3 medium. The plate was inserted into the Zebrabox (Viewpoint, France) allowing the fish to readapt to the equipment light conditions before subjected to seven Light-Dark cycles (including the light adaptation step) during the motion tracking. Dark duration was set to 1 second and light duration was set to 30 seconds. The overall activity of the larvae was recorded in one second-bins for a total duration of 3 minutes and 7 seconds following light-adaptation. Unresponsive larvae were defined as larvae moving less or equal to 0.1 mm per second during light to dark switches. For OMR, 9 dpf larvae were carefully transferred to a 10 well OMR plate (Viewpoint, France) with E3 medium. Adaptation to a white background (bottom screen) and lighting conditions was done for at least 30 minutes inside the Zebrabox (Viewpoint, France). Subsequently the background was changed into moving black horizontal stripes on a white background (10 stripes, 30 rpm, 25 fps) and fish locomotion was recorded for 5 minutes in 5 second bins. Swim speed was recorded in four categories: Fast (>8 mm/s), intermediate (3-8 mm/s) and slow (<3 mm/s) and no activity (no detection of movement). For all experiments light adaptation inside the Zebrabox and “Light periods” were set to 12 % power (Top Light) and “Dark periods” to 0 %, with an infrared illumination and camera recording at 60 frames per second. The detection sensitivity level was set to 21 and minimum size to 3, which focuses the movement detection on the head of each larva and ignores movement of smaller particles.

### Quantification and data analysis

Quantification of MGCs number and soma area in IF paraffin tissue sections was done blindly using the Cell Counter plugin in ImageJ (Fiji) and by randomly selecting six MGCs per section for manual area measurements. Cell of the GCL was counted from sections located near the middle of the eye using the Cell Counter plugin in ImageJ (Fiji), while the cell number for the other layers was estimated based on area measurements. BrdU^+^ and BLBP^+^ area measurements and PCNA^+^ cell counts in the dorsal and ventral CMZ were quantified from optical sections in the middle of the eye. ZPR3^+^ cell number, signal volume and fluorescence intensity were quantified in IF sections or whole eyes samples by extracting statistical data from spot or surface function in Imaris (Bitplane, UK). IF data analysis of proliferative MGCs was performed on 3D reconstructions of whole eye samples from BrdU labelled larvae using Imaris (Bitplane, UK). MGCs were counted as proliferative when double labelled with Alexa647 (BrdU) and RedX (BLBP) signal. In whole eyes of juvenile and adult ZPR3^+^ volume estimation was done on 3D reconstructions using Imaris (Bitplane, UK). Thickness for each layer in the adult eye was estimated as the mean of three measurements in fixed ROIs positioned dorsal, posterior and ventral to the lens in haematoxylin and eosin-stained sections of 7 mpf eyes. Data analysis from IF or haematoxylin and eosin-stained paraffin sections, was performed on 3-4 consecutive or near-consecutive sections for each fish (at least 3 fish). For IF sections of the adult fish eye, a 700 x 700 µm ROI near the lens for each section was used for data analysis. For whole eye sampling, n=3-5 eyes from 3-5 fish (interval thickness: 0.2-0.5 μm) or 12 consecutive optical sections for each genotype (3 fish; interval thickness: 5.0 μm) was used for data analysis. The pituitary macular region in 3D reconstructed eyes in the figures was estimated based on human eye anatomy^79^, at a fixed angle (17 degrees) with a distance of 2.5 times the optic nerve diameter from the optic nerve (14 dpf^+/+;+/-;-/-^: 63.4, 59.9 and 60.6 μm; 9 mpf^+/+;+/-;-/-^: 716.7, 733.8 and 682.6 μm).

## Data Availability

Individual genome sequencing data cannot be shared due to concerns over patient privacy. Other data generated or analyzed during this study are included in the main paper or its additional files.

## AUTHOR CONTRIBUTIONS

MVKS, TR, LAL and KG conceived the study. LAL, KG, STC and KM supervised experiments. MVKS, LAL and KG wrote the manuscript with input from all authors (MVKS, CR, SB, YA, LK, STC, KB-N, KM, TR, LAL and KG). CR, LK and TR performed clinical analyses. CJ and KG performed genetic analysis. MVKS, SB and YA performed zebrafish experiments and analyses. KM performed immunohistochemistry on fetal human sections.

## ACKNOWLEDGEMENTS

We are grateful to the patients and their family members for agreeing to participate in this study. We would like to thank Pernille Froh, Ha Nguyen and Pia Skovgaard for expert technical assistance. We acknowledge the Core Facility for Integrated Microscopy, Faculty of Health and Medical Sciences, University of Copenhagen. The study was supported by grants from the Velux Foundation, the Novo Nordisk Foundation and the Carlsberg Foundation.

## SUPLEMENTAL FIGURES & FIGURE LEGENDS

**Figure S1.**
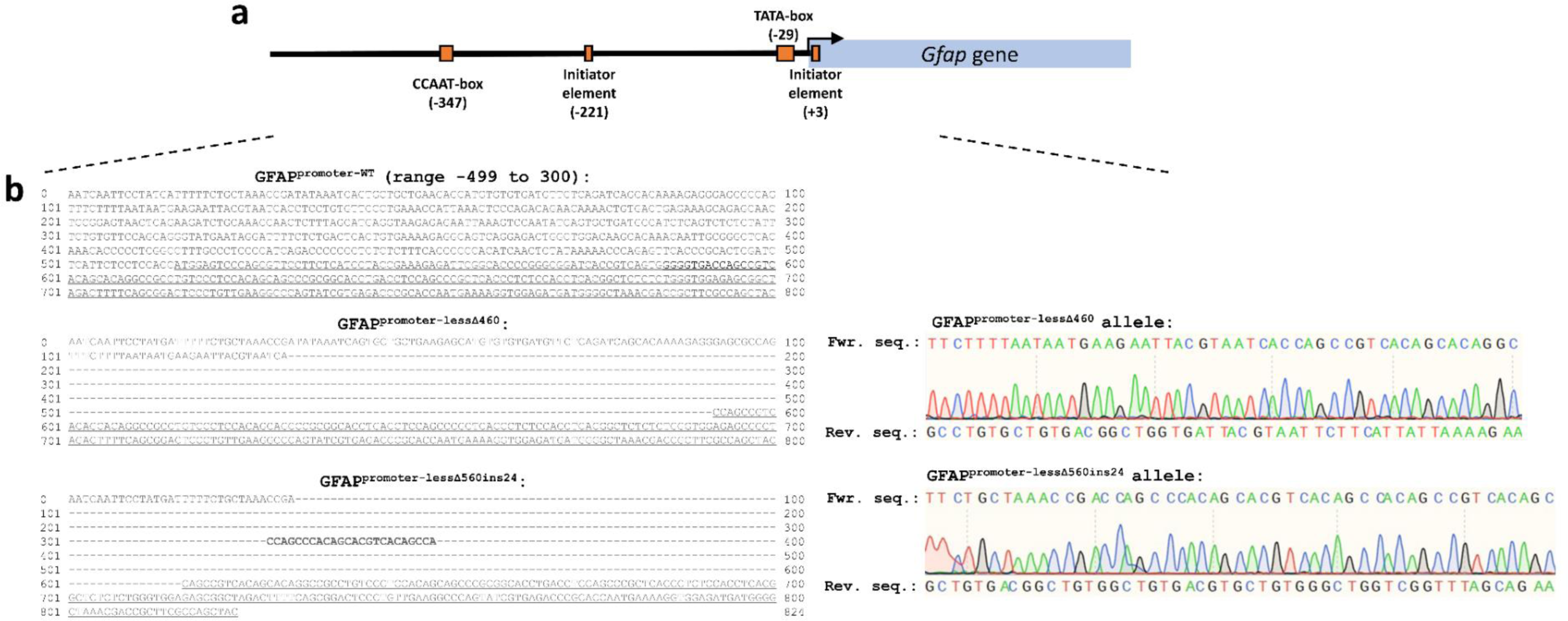
Analysis of the zebrafish gfap promoter and sequence of gfap promoter-less mutants. **a.** Diagram of the *Danio rerio* (zebrafish) *gfap* promoter (ENSDARG00000025301) from -500 to +300 and result of *in silico* analysis of promoter motifs in the same region with a cut-off p-value of: 0.001. **b, left.** Comparison of the *gfap* promoter and coding sequence from -499 to +300 in WT, gfap^Δ4^^60^ and gfap^Δ560ins24^ promoter-less mutants. Underlined and bold text shows exon and insertion sequence, respectively. **b, right.** Chromatogram obtained with forward sequencing primer and the sequence obtained from the reverse sequencing primer in the affected regions of the same fish lines.

**Figure S2.**
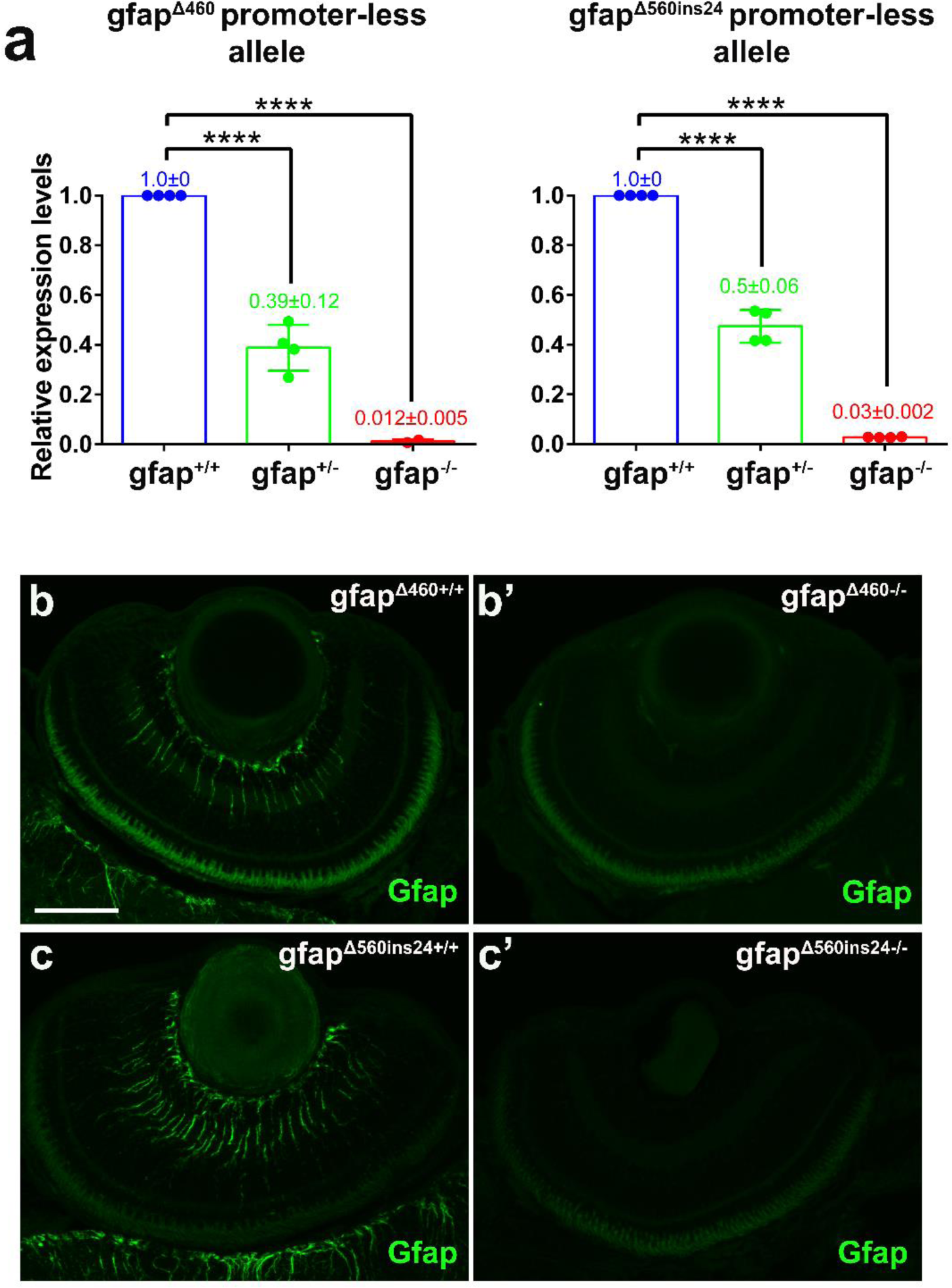
Silencing gfap promoter in promoter-less zebrafish mutants result in no detectable Gfap protein. **a.** Relative expression of zebrafish *gfap* mRNA in 3 dpf gfap^Δ460^ and gfap^Δ560ins24^ promoter-less mutants and WT siblings quantified by qRT-PCR (n=2-4 for each genotype). b-c’. IF maximum projection of Gfap^+^ MGC endfeet and processes, as well as astrocytes in 6 dpf gfap^Δ460^ (b,b’) and gfap^Δ560ins24^ (c,c’) WT and mutant retina. Scalebar, 50 μm.

**Figure S3.**
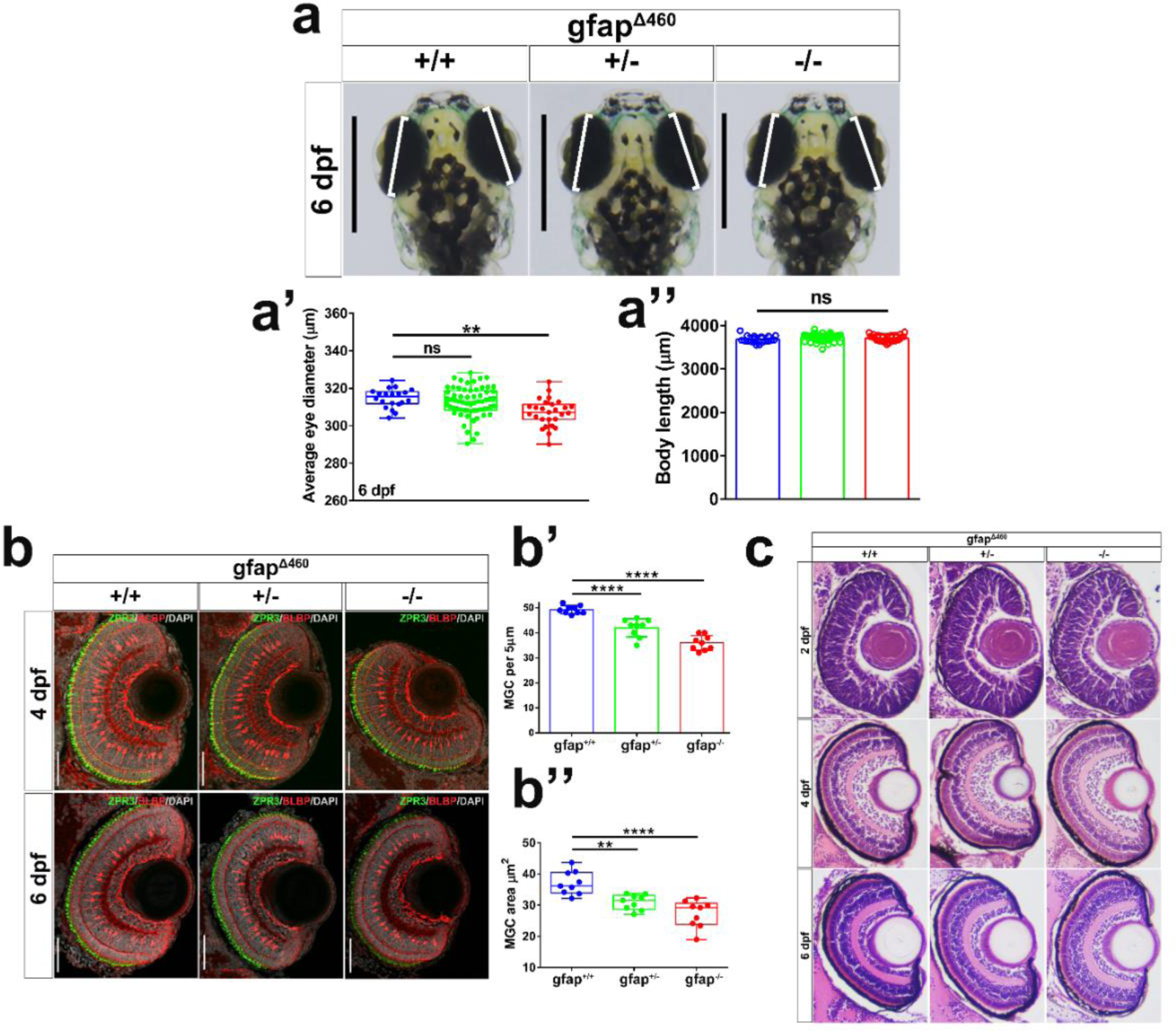
Characterization of gfap^Δ460^ promoter-less mutants. **a.** Dorsal view images of 6 dpf larvae head of uninjected WT and mutant siblings. White bars indicate an approximation of the major axis diameter for comparison, scale bar: 50 μm. **a’**. Measured mean diameter of both eyes for uninjected 6 dpf larvae (n_WT_= 20, n_+/-_=64, n_-/-_=26). **a’’**. Bar graph of measured larva body length of the fish shown in a’. **b.** IF maximum projection of BLBP^+^ MGCs and ZPR3^+^ rods and double cone outer segment in the OS of 4 dpf and 6dpf retinas from mutant and WT siblings. Scale bar: 50 μm. **b’.** Bar plot of the MGC count per 5μm section (n=9 for each genotype). **b’.** Quantification of soma area of MGCs in 4 and 6 dpf retinas from the same group of fish as shown in (b) (n=9 for each genotype). **c.** Hematoxylin and eosin staining of WT and mutant siblings eye sections at different stages: 2, 4 and 6 dpf. The data are presented as mean±sd. Asterisks indicate P-values obtained by using One-way ANOVA test: *: P < 0.05, **: P < 0.01, ****: P < 0.0001, ns: not significant.

**Figure S4.**
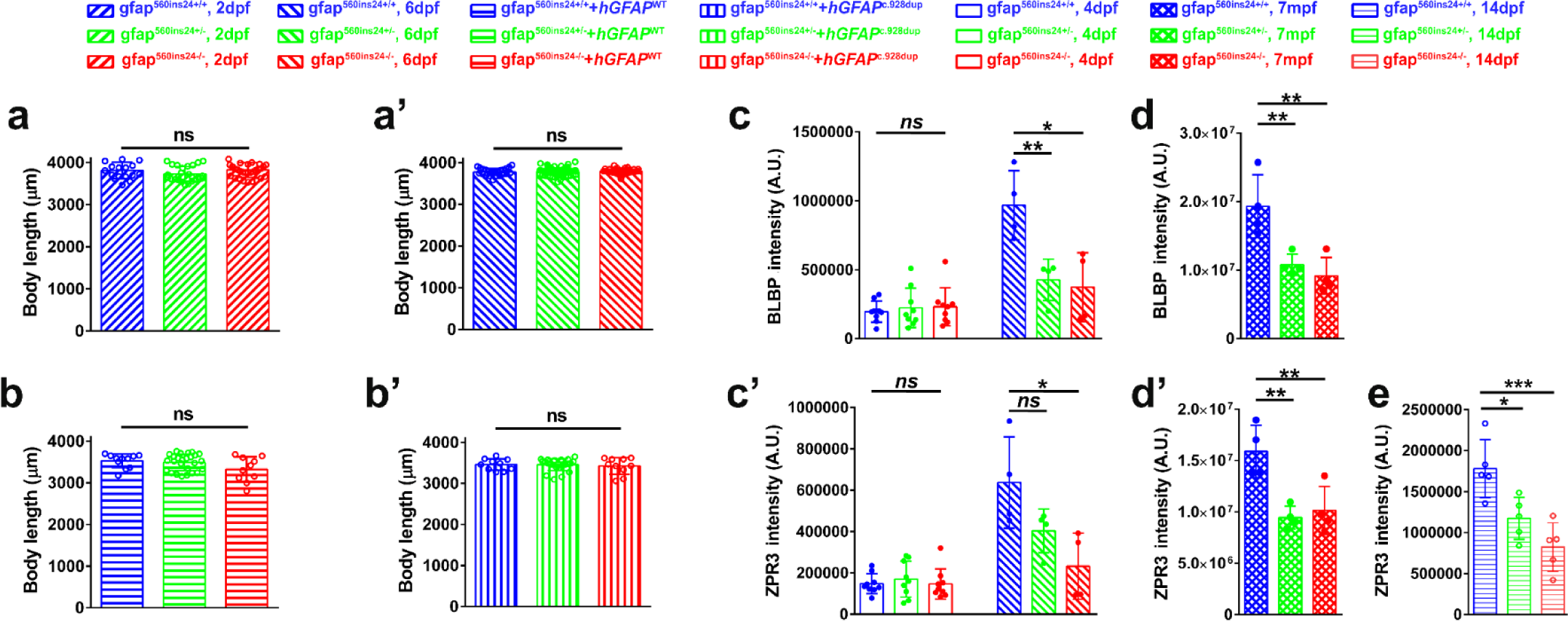
Additional data for gfap^Δ560ins24^ promoter-less mutants. **a-b’.** Bar graph of measured larva body length of uninjected 2 dpf (a), 6 dpf uninjected (a’) and injected with100pg WT gfap mRNA (b) or c.928dup mRNA (b’) in mutants and WT siblings group shown in Figure 4. **c-d’**. Bar graphs of BLBP (c, d) and ZPR3 (c’,d’) signal intensities measured in the ONL and OS layer of the eyes from 4 and 6 dpf as well as 7 mpf mutants and their WT siblings fish (n=4 for each genotype). **e.** Bar graph of ZPR3 signal intensities measured in eyes from 14 dpf mutants and WT siblings fish (n=5 for each genotype).

**Figure S5.**
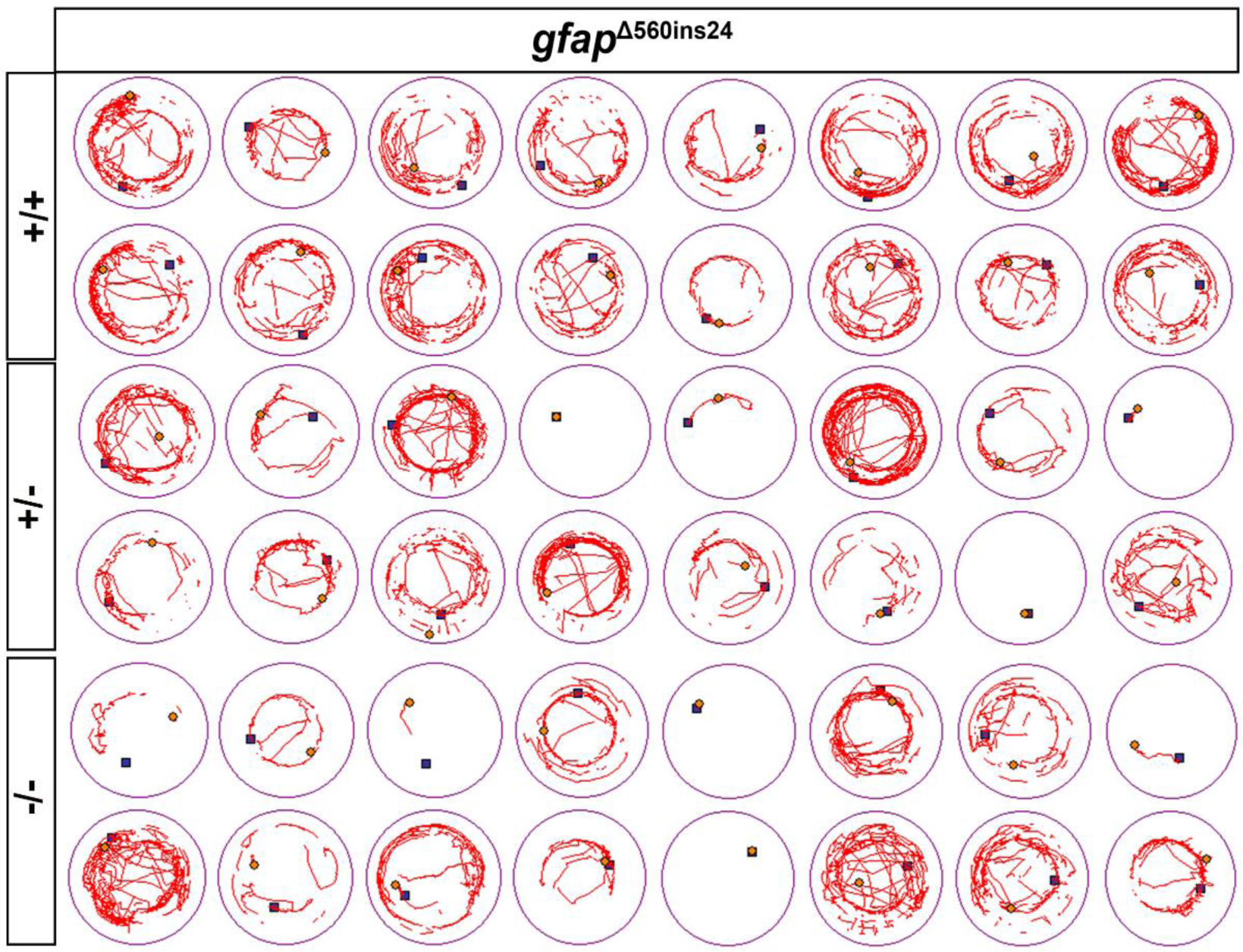
Startle response locomotion of *gfap*^Δ560ins24^ promoter-less mutants. Representative locomotion of sixteen 7 dpf gfap^Δ560ins24^ promoter-less mutants and their WT siblings. Red lines indicate the movement of the individual larva in each well during the time recorded.

**Figure S6.**
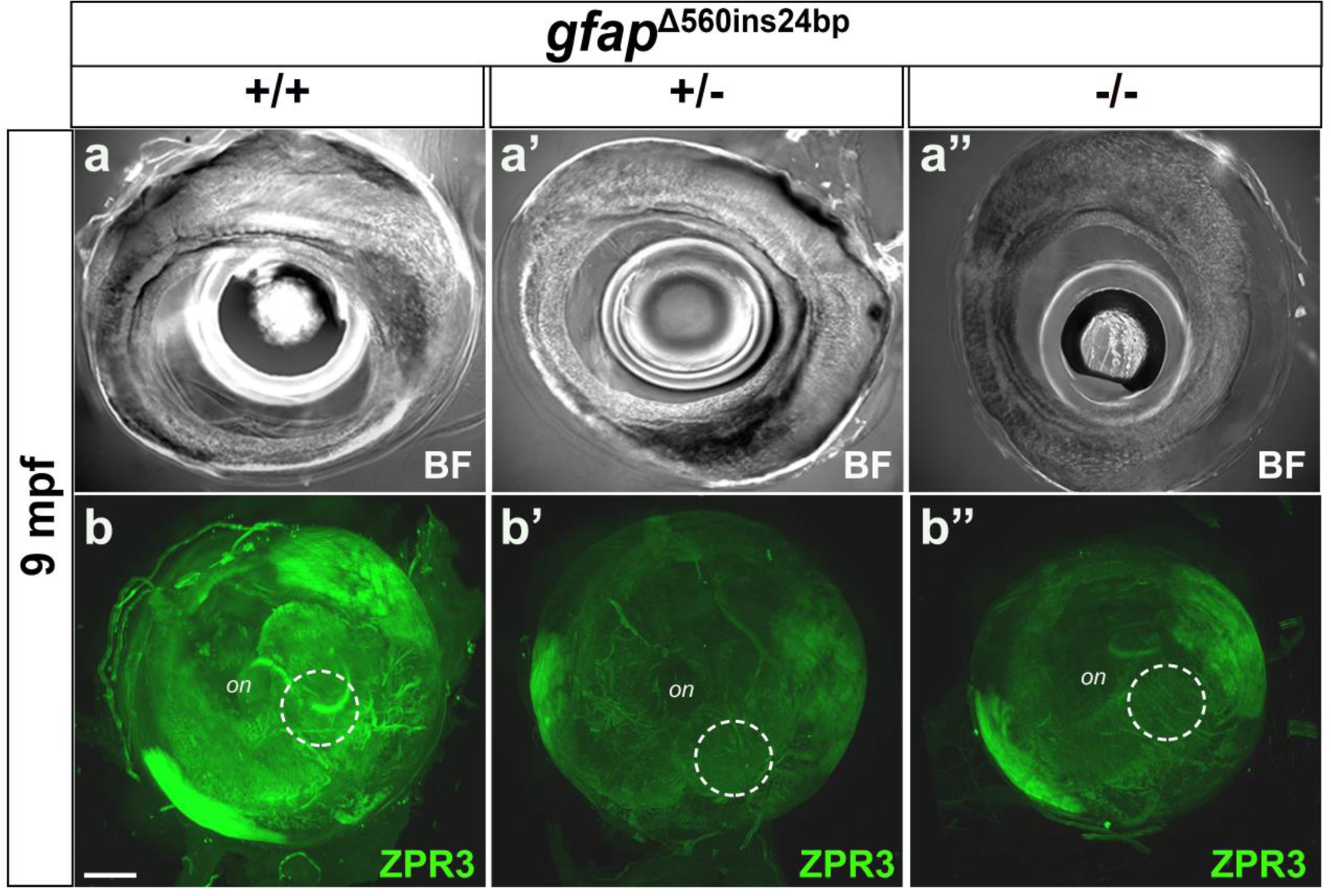
Rods and double cone outer segment of *gfap*^Δ560ins24^ promoter-less adult fish eyes. **a-a’’.** Brightfield pictures of adult eyes from 9 mpf WT siblings (a) and gfap^Δ560ins24^ promoter-less mutant fish (a’,a’’) (not to scale). **b-b’’.** 3D reconstruction of the retina stained for ZPR3^+^ rods and double cones in the same group of fish eyes (posterior view), scale bar 300 μm. White circle highlights the pituitary macular region of the eye.

## SUPLEMENTAL TABLES

**Table S1.** Clinical and genetic data on family. ^a^ GFAP NC_000017.11, NM_002055.5 (GRCh38). WT: normal sequence in GFAP position c.928. ^b^ No retinal dragging was observed in any of the affected individuals. BCVA: best corrected visual acuity. NA: not applied. RE: right eye; LE: left eye.

| Position in pedigree | IV-6 | V-1 | V-3 | V-7 |
| --- | --- | --- | --- | --- |
| Genotype GFAP<sup>a</sup> | c.[928dup];[=] | c.[928dup];[=] | c.[928dup];[=] | c.[928dup];[=] |
| BCVA |  | 1982:<br>RE: 1,25 (+0,75 -2,0 x 80°)<br>LE: 1,25 (+0,75 -1,75 x 150°) | 1982:<br>RE: 0,3 (+3,5 -2,25 x 80°)<br>LE: 0,25 (+4,5 -1,0 x 145°)<br>2003:<br>RE: 0,4 (+2,75 -2,25 x 70°)<br>LE: 0,3 (+3,25 -1,0 x 90°) | 1981:<br>RE:0,25 (+0,75)<br>LE: 0,16 (-1,0 -2,0 x 0°)<br>2005:<br>RE: 0,16 (+1,5 -2,25 x 170°)<br>LE: 0,16 (+2,0 -2,75 x 5°) |
| Fundoscopy<sup>b</sup> | Normal looking optic nerve head, retina, and vascular system | Pre- and peripapillary pellucid veil, normal optic nerve head, macular region and normal vessels | Massive pre-and peripapillary glial mass with sheeting of arterioles extending along the arcades. Distinct macular RPE atrophy in both macular regions | Pellucid pre- and peripapillary glial mass, massive central and peripheral vascular sheeting with a few splint hemorrhages. Diffuse RPE atrophy in the macular regions with pigment clumping. |
| Goldmann perimetry | Normal | Normal | Central scotoma both eyes | Central scotoma both eyes |
| Colour vision | NA | NA | Normal | Normal |
| Fullfield ERG | NA | NA | Normal | Normal |
| OCT |  | Abnormal morphology of optic disc. No signs of drusen. Extension of the epiretinal glial mass to macular region. Disorganized inner retinal layer | Abnormal morphology of optic disc. No signs of drusen. Extension of the epiretinal glial mass to macular region. Disorganized inner retinal layer. Atrophic retina in foveal region. | Abnormal morphology of optic disc. No signs of drusen. Extension of the epiretinal glial mass to macular region. Disorganized inner retinal layer. Atrophic retina in foveal region. |
| Fluorescein angiography 1974/2018 |  | NA | NA | Vascularized epipapillary glial mass |
| Systemic signs |  | None | None | None |

**Table S2.** Rare variants shared by affected family members. Three affected individuals (V-1, V-3 and V-7) and one unaffected individual (IV-4) was analyzed using whole genome sequencing. Rare variants present in all three affected individuals, but not in the unaffected individual is shown.

| Gene | Chr location (GRCh37) | Transcript | Protein | Population frequency <sup>a</sup> | Consequence | Classification <sup>b</sup> | Disease association of gene |
| --- | --- | --- | --- | --- | --- | --- | --- |
| *PEMT* | 17:17494884 | NM_148172.2:<br>c.57T>C | NP_680477.1:<br>p.Pro19= | 7.15e-04 | Synonymous | n/a | n/a |
| *OR10H2* | 19:15839201 | NM_013939.2:<br>c.348C>T | NP_039227.1:<br>p.Thr116= | 1.79e-04 | Synonymous | n/a | n/a |
| *GAS2L2* | 17:34074257 | NM_139285.3:<br>c.863C>T | NP_644814.1:<br>p.Pro288Leu | 8.77e-04 | Missense | VUS (PM2_sup, BP4_mod) | Biallelic variants associated with primary ciliary dyskinesia (MIM #618449)<sup>18</sup> |
| *SOST* | 17:41836027 | NM_025237.2:<br>c.83C>T | NP_079513.1:<br>p.Ala28Val | 2.73e-05 | Missense | VUS (BP4_sup) | Monoallelic variants associated with Craniodiaphyseal dysplasia (MIM #122860)<sup>21</sup>. Biallelic variants associated with Sclerosteosis 1 (MIM #269500)<sup>22</sup> |
| *GFAP* | 17:42988803 | NM_001242376.1:<br>c.928dupA | NP_001229305.1:<br>p.Met310Asnfs | n/a | Frameshift | Likely pathogenic (PVS1_mod, PM2_sup, PP1_strong) | Monoallelic variants associated with Alexander disease (MIM # 203450). |

<sup>a</sup> Frequency in gnomAD v.4.0.0 (gnomad.broadinstitute.org). <sup>b</sup> Classification according to ACMG guidelines<sup>19</sup> and ClinGenome webpage<sup>20</sup>

## REFERENCES

1. Messing, A. & Brenner, M. GFAP at 50. ASN Neuro 12, (2020).

2. Tao, C. & Zhang, X. Development of astrocytes in the vertebrate eye. Dev Dyn 243, 1501–10 (2014).

3. Holst, C. B., Brøchner, C. B., Vitting-Seerup, K. & Møllgård, K. Astrogliogenesis in human fetal brain: complex spatiotemporal immunoreactivity patterns of GFAP, S100, AQP4 and YKL-40. J Anat 235, 590–615 (2019).

4. Sarthy, P. V., Fu, M. & Huang, J. Developmental Expression of the Glial Fibrillary Acidic Protein (GFAP) Gene in the Mouse Retina. Cellular and Molecular Neurobiology vol. 11 (1991).

5. Sánchez-Farías, N. & Candal, E. Identification of radial glia progenitors in the developing and adult retina of sharks. Front Neuroanat 10, (2016).

6. Julian, D., Ennis, K. & Korenbrot, J. I. Birth and fate of proliferative cells in the inner nuclear layer of the mature fish retina. Journal of Comparative Neurology 394, 271– 282 (1998).

7. Otteson, D. C., D’Costa, A. R. & Hitchcock, P. F. Putative stem cells and the lineage of rod photoreceptors in the mature retina of the goldfish. Dev Biol 232, 62–76 (2001).

8. Nelson, S. M., Frey, R. A., Wardwell, S. L. & Stenkamp, D. L. The developmental sequence of gene expression within the rod photoreceptor lineage in embryonic zebrafish. Developmental Dynamics 237, 2903–2917 (2008).

9. Mack, A. F., Papanikolaou, D. & Lillo, C. Investigation of the migration path for new rod photoreceptors in the adult cichlid fish retina. Exp Neurol 184, 90–96 (2003).

10. Fischer, A. J. & Reh, T. A. Müller glia are a potential source of neural regeneration in the postnatal chicken retina. Nat Neurosci 4, 247–52 (2001).

11. Brenner, M. et al. Mutations in GFAP, encoding glial fibrillary acidic protein, are associated with Alexander disease. Nat Genet 27, 117–20 (2001).

12. Hagemann, T. L. Alexander disease: models, mechanisms, and medicine. Current Opinion in Neurobiology vol. 72 140–147 Preprint at 10.1016/j.conb.2021.10.002 (2022).

13. Messing, A. Alexander disease. in Handbook of Clinical Neurology vol. 148 693–700 (Elsevier B.V., 2018).

14. Nam, T. S. et al. Identification of a novel nonsense mutation in the rod domain of GFAP that is associated with Alexander disease. European Journal of Human Genetics 23, 72–78 (2015).

15. Messing, A. Refining the concept of GFAP toxicity in Alexander disease. Journal of Neurodevelopmental Disorders vol. 11 Preprint at 10.1186/s11689-019-9290-0 (2019).

16. Hagemann, T. L. et al. Antisense suppression of glial fibrillary acidic protein as a treatment for Alexander disease. Ann Neurol 83, 27–39 (2018).

17. Hagemann, T. L., et al. Antisense Therapy in a Rat Model of Alexander Disease Reverses GFAP Pathology, White Matter Deficits, and Motor Impairment. Sci. Transl. Med vol. 13 https://www.science.org (2021).

18. Bustamante-Marin, X. M. et al. Lack of GAS2L2 Causes PCD by Impairing Cilia Orientation and Mucociliary Clearance. Am J Hum Genet 104, 229–245 (2019).

19. Richards, S. et al. Standards and guidelines for the interpretation of sequence variants: a joint consensus recommendation of the American College of Medical Genetics and Genomics and the Association for Molecular Pathology. Genet Med 17, 405–24 (2015).

20. ClinGen The Clinical Genome Resource. Heidi L. Rehm, Ph.D., Jonathan S. Berg, M.D., Ph.D., Lisa D. Brooks, Ph.D., Carlos D. Bustamante, Ph.D., James P. Evans, M.D., Ph.D., Melissa J. Landrum, Ph.D., David H. Ledbetter, Ph.D., Donna R. Maglott, Ph.D., Christa Lese Martin, Ph.D., Robert L. Nussbaum, M.D., Sharon E. Plon, M.D., Ph.D., Erin M. Ramos, Ph.D., Stephen T. Sherry, Ph.D., and Michael S. Watson, Ph.D., for ClinGen. N Engl J Med 2015; 372:2235–2242 June 4, 2015 DOI: 10.1056/NEJMsr1406261.. https://clinicalgenome.org/working-groups/sequence-variant-interpretation/.

21. Kim, S. J. et al. Identification of signal peptide domain SOST mutations in autosomal dominant craniodiaphyseal dysplasia. Hum Genet 129, 497–502 (2011).

22. Brunkow, M. E. et al. Bone dysplasia sclerosteosis results from loss of the SOST gene product, a novel cystine knot-containing protein. Am J Hum Genet 68, 577–589 (2001).

23. Lee, S. H. et al. Aggregation-prone GFAP mutation in Alexander disease validated using a zebrafish model. BMC Neurol 17, (2017).

24. El-Brolosy, M. A. et al. Genetic compensation triggered by mutant mRNA degradation. Nature 568, 193–197 (2019).

25. Bernardos, R. L. & Raymond, P. A. GFAP transgenic zebrafish. Gene Expression Patterns 6, 1007–1013 (2006).

26. Fischer, A. J., Bosse, J. L. & El-Hodiri, H. M. The ciliary marginal zone (CMZ) in development and regeneration of the vertebrate eye. Exp Eye Res 116, 199–204 (2013).

27. Anthony, T. E., Mason, H. A., Gridley, T., Fishell, G. & Heintz, N. Brain lipid-binding protein is a direct target of Notch signaling in radial glial cells. Genes Dev 19, 1028– 1033 (2005).

28. Raymond, P. A., Barthel, L. K., Bernardos, R. L. & Perkowski, J. J. Molecular characterization of retinal stem cells and their niches in adult zebrafish. BMC Dev Biol 6, 36 (2006).

29. Bernardos, R. L., Barthel, L. K., Meyers, J. R. & Raymond, P. A. Late-stage neuronal progenitors in the retina are radial Müller glia that function as retinal stem cells. Journal of Neuroscience 27, 7028–7040 (2007).

30. Yin, J. et al. The 1D4 antibody labels outer segments of long double cone but not rod photoreceptors in zebrafish. Invest Ophthalmol Vis Sci 53, 4943–4951 (2012).

31. Noel, N. C. L., MacDonald, I. M. & Allison, W. T. Zebrafish Models of Photoreceptor Dysfunction and Degeneration. Biomolecules 11, 1–33 (2021).

32. Zhu, M., Provis, J. M. & Penfold, P. L. The human hyaloid system: cellular phenotypes and inter-relationships. Exp Eye Res 68, 553–63 (1999).

33. Goldberg, M. F. Persistent fetal vasculature (PFV): an integrated interpretation of signs and symptoms associated with persistent hyperplastic primary vitreous (PHPV). LIV Edward Jackson Memorial Lecture. Am J Ophthalmol 124, 587–626 (1997).

34. Mehta, A., Singh, S. R., Dogra, M. & Ram, J. Persistent fetal vasculature - Clinical spectrum. Indian J Ophthalmol 66, 1860 (2018).

35. Sisk, R. A., Berrocal, A. M., Schefler, A. C., Dubovy, S. R. & Bauer, M. S. Epiretinal membranes indicate a severe phenotype of neurofibromatosis type 2. Retina 30, S51–8 (2010).

36. Xiao, H., Tong, Y., Zhu, Y. & Peng, M. Familial Exudative Vitreoretinopathy-Related Disease-Causing Genes and Norrin/β-Catenin Signal Pathway: Structure, Function, and Mutation Spectrums. J Ophthalmol 2019, 5782536 (2019).

37. Azuma, N., Hida, T. & Kohsaka, S. Hypovascular glial overgrowth from the optic nerve head in foetuses of 16 weeks gestation. Acta Ophthalmol 87, 355–7 (2009).

38. Gass, J. D. M. An unusual hamartoma of the pigment epithelium and retina simulating choroidal melanoma and retinoblastoma. 1973. Retina 23, 171–83; discussion 184-5 (2003).

39. Shields, C. L. et al. Combined hamartoma of the retina and retinal pigment epithelium in 77 consecutive patients visual outcome based on macular versus extramacular tumor location. Ophthalmology 115, 2246–2252.e3 (2008).

40. Hester CC, Imes RK, Lujan BJ & Fu AD. Autosomal dominant bilateral combined hamartoma of the retina and retinal pigment epithelium or a new familial optic nerve dysgenesis? Invest. Ophthalmol. Vis. Sci. 51, 3546 (2010).

41. Božanić, D., Bočina, I. & Saraga-Babić, M. Involvement of cytoskelatal proteins and growth factor receptors during development of the human eye. Anat Embryol (Berl) 211, 367–377 (2006).

42. Chu, Y., Hughes, S. & Chan-Ling, T. Differentiation and migration of astrocyte precursor cells (APCs) and astrocytes in human fetal retina: relevance to optic nerve coloboma. The FASEB Journal 15, 2013–2015 (2001).

43. Huxlin, K. R., Sefton, A. J. & Furby, J. H. The origin and development of retinal astrocytes in the mouse. J Neurocytol 21, 530–544 (1992).

44. Dorrell, M. I., Aguilar, E. & Friedlander, M. Retinal vascular development is mediated by endothelial filopodia, a preexisting astrocytic template and specific R-cadherin adhesion. Invest Ophthalmol Vis Sci 43, 3500–10 (2002).

45. Mellough, C. B. et al. An integrated transcriptional analysis of the developing human retina. Development (Cambridge) 146, (2019).

46. Wetts, R., Serbedzija, G. N. & Fraser, S. E. Cell lineage analysis reveals multipotent precursors in the ciliary margin of the frog retina. Dev Biol 136, 254–63 (1989).

47. Johns, P. R. Growth of the adult goldfish eye. III. Source of the new retinal cells. J Comp Neurol 176, 343–57 (1977).

48. Fischer, A. J. & Reh, T. A. Identification of a proliferating marginal zone of retinal progenitors in postnatal chickens. Dev Biol 220, 197–210 (2000).

49. Marcucci, F. et al. The Ciliary Margin Zone of the Mammalian Retina Generates Retinal Ganglion Cells. Cell Rep 17, 3153–3164 (2016).

50. Bélanger, M.-C., Robert, B. & Cayouette, M. Msx1-Positive Progenitors in the Retinal Ciliary Margin Give Rise to Both Neural and Non-neural Progenies in Mammals. Dev Cell 40, 137–150 (2017).

51. Bulirsch, L. M. et al. Spatial and temporal immunoreaction of nestin, CD44, collagen IX and GFAP in human retinal Müller cells in the developing fetal eye. Exp Eye Res 217, (2022).

52. Sugiyama, A. et al. Incidental diagnosis of an asymptomatic adult-onset Alexander disease by brain magnetic resonance imaging for preoperative evaluation. Journal of the Neurological Sciences vol. 354 131–132 Preprint at 10.1016/j.jns.2015.05.001 (2015).

53. Brenner, M. & Messing, A. A new mutation in GFAP widens the spectrum of Alexander disease. Eur J Hum Genet 23, 1–2 (2015).

54. Quinlan, R. A., Moir, R. D. & Stewart, M. Expression in Escherichia coli of fragments of glial fibrillary acidic protein: characterization, assembly properties and paracrystal formation. J Cell Sci 93 ( **Pt 1**), 71–83 (1989).

55. Chen, W. J. & Liem, R. K. The endless story of the glial fibrillary acidic protein. J Cell Sci 107 ( **Pt 8**), 2299–311 (1994).

56. Perng, M.-D. et al. Glial fibrillary acidic protein filaments can tolerate the incorporation of assembly-compromised GFAP-delta, but with consequences for filament organization and alphaB-crystallin association. Mol Biol Cell 19, 4521–33 (2008).

57. Flint, D. et al. Splice site, frameshift, and chimeric GFAP mutations in Alexander disease. Hum Mutat 33, 1141–1148 (2012).

58. van Asperen, J. V, Robe, P. A. J. T. & Hol, E. M. GFAP Alternative Splicing and the Relevance for Disease - A Focus on Diffuse Gliomas. ASN Neuro 14, 17590914221102064 (2022).

59. Maquat, L. E. Nonsense-mediated mRNA decay in mammals. J Cell Sci 118, 1773– 1776 (2005).

60. Gomi, H., et al. Mice Devoid of the Glial Fibrillary Acidic Protein Develop Normally and Are Susceptible to Scrapie Prions. Neuron vol. 14 (1995).

61. Mccall, M. A., et al. Targeted Deletion in Astrocyte Intermediate Filament (Gfap) Alters Neuronal Physiology (Long-Term Potentiation/Hippocampus/Optic Nerve/Homologous Recombination/Mouse). Neurobiology vol. 93 https://www.pnas.org (1996).

62. Pekny, M. et al. Mice lacking glial fibrillary acidic protein display astrocytes devoid of intermediate filaments but develop and reproduce normally. EMBO Journal 14, 1590– 1598 (1995).

63. Nawashiro, H., Messing, A., Azzam, N. & Brenner, M. Mice lacking GFAP are hypersensitive to traumatic cerebrospinal injury. Neuroreport 9, 1691–6 (1998).

64. Shibuki, K., Gomi, H. & Chen, L. Deficient Cerebellar Long-Term Depression, Impaired Eyeblink Conditioning, and Normal Motor Coordination in GFAP Mutant Mice. Neuron vol. 16 (1996).

65. Triolo, D. et al. Loss of glial fibrillary acidic protein (GFAP) impairs Schwann cell proliferation and delays nerve regeneration after damage. J Cell Sci 119, 3981–3993 (2006).

66. Kamphuis, W. et al. GFAP and vimentin deficiency alters gene expression in astrocytes and microglia in wild-type mice and changes the transcriptional response of reactive glia in mouse model for Alzheimer’s disease. Glia 63, 1036–1056 (2015).

67. Rossi, A. et al. Genetic compensation induced by deleterious mutations but not gene knockdowns. Nature 524, 230–233 (2015).

68. Fernald, R. D. Teleost vision: Seeing while growing. Journal of Experimental Zoology 256, 167–180 (1990).

69. MacDonald, R. B. et al. Müller glia provide essential tensile strength to the developing retina. Journal of Cell Biology 210, 1075–1083 (2015).

70. Verardo, M. R. et al. Abnormal reactivity of muller cells after retinal detachment in mice deficient in GFAP and vimentin. Invest Ophthalmol Vis Sci 49, 3659–65 (2008).

71. Shen, W. et al. Conditional Müller cell ablation causes independent neuronal and vascular pathologies in a novel transgenic model. J Neurosci 32, 15715–27 (2012).

72. Byrne, L. C. et al. AAV-Mediated, Optogenetic Ablation of Müller Glia Leads to Structural and Functional Changes in the Mouse Retina. PLoS One 8, e76075 (2013).

73. Maggs, A. & Scholes, J. Glial domains and nerve fiber patterns in the fish retinotectal pathway. J Neurosci 6, 424–38 (1986).

74. Conrad, M., Lemb, K., Schubert, T. & Markl, J. Biochemical identification and tissue-specific expression patterns of keratins in the zebrafish Danio rerio. Cell Tissue Res 293, 195–205 (1998).

75. van Cruchten, S. et al. Pre- and Postnatal Development of the Eye: A Species Comparison. Birth Defects Res 109, 1540–1567 (2017).

76. Harris, W. A. & Perron, M. Molecular recapitulation: the growth of the vertebrate retina. Int J Dev Biol 42, 299–304 (1998).

77. Pende, M. et al. A versatile depigmentation, clearing, and labeling method for exploring nervous system diversity. Sci Adv 6, eaba0365 (2020).

78. Kimura, T. et al. Guanine crystals regulated by chitin-based honeycomb frameworks for tunable structural colors of sapphirinid copepod, Sapphirina nigromaculata. Sci Rep 10, (2020).

79. Kolb, H. Simple Anatomy of the Retina. Webvision: The Organization of the Retina and Visual System (1995).

